# Quantification of Brain Functional Connectivity Deviations in Individuals: A Scoping Review of Functional MRI Studies

**DOI:** 10.1101/2024.12.12.24318915

**Authors:** Artur Toloknieiev, Dmytro Voitsekhivskyi, Hlib Kholodkov, Roman Lvovich, Petro Matiushko, Daria Rekretiuk, Andrii Dikhtiar, Antonii Viter, Volodymyr Pokras, Stephan Wunderlich, Sophia Stoecklein

## Abstract

Functional connectivity magnetic resonance imaging (fcMRI) is a well-established technique for studying brain networks in both healthy and diseased individuals. However, no fcMRI-based biomarker has yet achieved clinical relevance. To establish better understanding of the state of the art in quantifying abnormal connectivity in comparison to a reference distribution, for potential use in individual patients, we have conducted a scoping review over 5672 entries from the last 10 years. We have located five publications proposing metrics of abnormal connectivity quantification, reported these metrics, formalized their computing methods, assessed their technology readiness and estimated their computational efficiency. Building upon our findings, we have discussed the metrics’ lesion data handling, region of interest level of detail and potential clinical use cases. We also proposed methodical and computational strategies for improvement of current and emerging abnormality quantification metrics in fcMRI research.

## 1. Introduction

### 1.1 Resting-state functional connectivity MRI and its potential as a basis for clinically feasible diagnostic approaches

Functional connectivity magnetic resonance imaging (fcMRI), first used for connectivity analysis in humans by Biswal et al., [1] and based on the blood oxygen level dependent (BOLD) signal [2, 3, 4], is widely regarded as a valuable imaging method for the inquiry into connectivity in human [5, 6] and non-human [7] brain research alike. With the scientific community increasingly reconceptualizing neurodegenerative [8, 9], psychiatric [10] and neuro-oncological [11, 12, 13] disorders as “network disorders”, fcMRI-based biomarkers that quantify abnormal connectivity in relation to the distribution in a healthy reference sample may pave a way for individualized connectivity metrics suited for validation and application in clinical diagnostics.

FcMRI-based methods have been previously utilized in a wide variety of contexts. For instance, fcMRI was employed to guide repetitive transcranial magnetic stimulation (TMS) in depression [14], Parkinson disease [15] and post-stroke aphasia [16], and can be helpful for informing presurgical planning in brain tumor patients [17]. Likewise, fcMRI-derived biomarkers can be used to aid prognostication in neuro-oncological patients [13], as an association between overall survival and tumor connectivity to distant brain regions has been reported [18]. Moreover, reports of correlation between tau accumulation rates and connectivity strength to tau epicenters [8], as well as of spatial association of functional connectivity changes with microglial activation in A*β*- and tauopathies such as Alzheimer’s disease [9] suggest high utility of fcMRI-based biomarkers in diagnosis and prognostication of neurodegeneration. Lastly, fcMRI-based metrics have been used for therapy control and adverse effect management in hemato-oncological conditions [19], and thus have a potential to improve adverse effect management in systemic therapies. These examples illustrate only a subset of potential fcMRI applications, suggesting broader clinical applicability.

When aiming at clinical applicability, resting-state fcMRI (rs-fcMRI) exhibits various advantages over task-based fMRI. Firstly, it is easier to implement in routine clinical scanning as it does not require additional equipment, such as MRI-compatible monitors. Secondly, it is independent of the patient’s language proficiency and ability to follow task-paradigms. Finally, it provides a holistic assessment of the entire brain’s functional connectivity architecture, rather than assessing specific task domains or networks. We have therefore focused this scoping review on the rs-fcMRI literature.

### 1.2 Current Challenges in the Adoption of rs-fcMRI-derived Metrics

To date, no fcMRI biomarker has achieved clinical relevance. This can be attributed to limited interpretability and generalizability of rs-fcMRI data stemming from a set of factors, most notably biological variance, technical variance and a distinct lack of established normative ranges for functional connectivity in humans. A short overview is given below.

#### 1.2.1 Biological Variance

The first significant factor of biological variance consists of intra-individual variance in biological dynamics. For example, respiratory cycle and cardiovascular variables such as diastolic blood pressure influence the effective connectivity variance between and within resting-state networks [20]. Additionally, circadian rhythms, menstrual cycle, substance influence, emotional state [21] and self-generated thoughts (“mind-wandering”) [22] have been described as sources of intra-subject variance in rs-fcMRI experiments.

#### 1.2.2 Technical Variance

The factor of technical variance is a complex one. It consists of variance of data quality and is defined by scanner-, participant-, acquisition protocol-and processing-related influences. The scanner itself constitutes the first major source of potential variabilities, with field strength, scanner vendor and precise scanner element and upgrade composition being described as relevant confounders [23], all leading to a variable spatial distribution of the signal-to-noise and contrast-to-noise ratios of the BOLD signal across the brain, which in turn significantly influence connectivity metrics [24, 25]. Field inhomogeneities and MRI artifacts such as ghosting [26] or drift artifacts [27] and thermal noise [28, 29] have been shown to considerably influence the fcMRI data quality. Moreover, recent studies have demonstrated that vendor-specific slice- and volume-censoring techniques can also bias voxel-level connectivity estimates [30]. The influence of acquisition protocol aspects constitutes another major source of variability in quality of data for analytical and statistical purposes. For instance, the test-retest reliability of fcMRI experiments depends on both, the session scan duration and the number of sessions [31, 25], while dynamic functional connectivity test-retest reliability was demonstrated to depend significantly on the choice of sliding window size [32].

The participant is another considerable source of technical variance. Particularly, head motion has long been known to drastically impact functional connectivity measures [33], and has been related to physiological, behavioral and demographic measures in Human Connectome Project (HCP) data [34]. Patient data are especially prone to motion artifacts and might require denoising strategies that are tailored to the respective patient cohort [35].

Lastly, data processing pipelines are known to significantly influence data quality. FcMRI preprocessing operations are not commutative, as their order is capable of reintroducing artifacts into the data [36], and the specifics of the dataset itself (which are, in turn, determined by scanner, participant and protocol) strongly determine its compatibility with individual sequences of pre-processing operations [37]. Similarly, concatenation of data for purposes of data maximization as part of data preprocessing or various data wrangling operations may both improve reliability and affect it adversely, if done without sufficient caution [38].

#### 1.2.3 Absence of Normative Reference Distributions

One key barrier to clinical translation of fcMRI-based imaging markers is the lack of standardized, accessible reference distributions for human functional connectivity - despite large available datasets [39, 40, 41, 42]. As in other diagnostic fields, individual connectivity measurements must be benchmarked against a normative distribution to determine deviations from typical brain functional architecture. Inspired by successful normalization of imaging-derived markers in other modalities [43, 44, 45], establishing an fcMRI-based normative framework to both delineate “healthy” connectivity and quantify patient-specific divergence is clearly warranted. From this unmet need, two key assertions emerge.

Firstly, there exists an apparent unmet medical need for validated and clinically implemented fcMRI-based connectivity abnormality metrics that satisfy the criteria of relationality and countability. Herein, a relational metric may be defined as a metric that relies on a control cohort sufficiently representative of the target individual, allowing to establish a normative model of connectivity that compares a given individual to a distribution of controls and quantifies the discrepancy, while a countable metric may be defined as an interval or rational metric that can be used as grounds for grading or comparison. Secondly, there is minimal study coverage pertaining to the introduction and validation of such metrics, which limits current insight into individualized abnormality detection in functional connectivity.

An initial step towards addressing the question of normative modelling in fcMRI consists in a scoping review of fcMRI-based metrics of connectivity abnormality, the results of which we present here. Within the scope of this paper, we review, analyze and formalize the connectivity abnormality metrics yielded by our search, explore the degree of their refinement, determine their readiness for clinical validation and estimate their computational complexities. Moreover, we discuss the metrics’ lesion data handling, region of interest level of detail (LOD) and the potential clinical use cases. We also propose methodical and computational strategies for improvement of current and emerging abnormality quantification metrics.

## 2 Methods

### 2.1 Overall Protocol

We have conducted our review in adherence to the general framework of scoping reviews proposed by Arksey and O’Malley [46] and refined by Levac et al. [47]. We reported our results in compliance with the Preferred Reporting Items for Systematic reviews and Meta-Analyses (PRISMA) extension for scoping reviews (PRISMA-ScR) [48]. The PRISMA-ScR compliance checklist can be accessed in the Supplementary Materials.

### 2.2 Review Objectives

Within the scope of this review, we intended to determine (1) whether there exists a way to quantify the deviation of functional connectivity in an individual patient from the general population, (2) whether it is validated to guarantee sufficient technology readiness and clinical utility and (3) whether it satisfies the criteria of relationality and countability outlined in the introduction. In pursuit of this objective, we have reviewed the state-of-the-art (SOTA) of personalized detection of connectivity abnormalities in brain fcMRI, analyzed the results, systematized them, and reported our findings.

### 2.3 Information Sources, Search Strategy, Data Acquisition and Handling

We have leveraged the Google Scholar database for our search. We set the query year range at the years 2014-2024 and employed Publish or Perish 8.10.4612.8838 [49] to automate our query. We searched in 1-year batches to yield the most entries and circumvent the internal limit of 1000 entries per query. We input the following search request: “fMRI connectivity connectome abnormality detection anomaly map deviation individual reference metric.” All data was aggregated using pandas 2.1.1 [50] and NumPy 1.23.5 [51], exported as CSV, and uploaded for subsequent group review on a secure team space in Notion [52]. Using Notion’s integrated tools and functions, we removed damaged or empty entries. The remaining entries have been subjected to screening and eligibility assessment (see below).

### 2.4 Study Screening and Selection

We employed a PRISMA compliant, 2-phase screening and eligibility selection strategy. During the screening phase, we excluded sources that (1) did not report research based on fcMRI and did not use BOLD signal, (2) reported experiments on participants under 18 years of age, (3) did not have a healthy reference cohort against which the patients would be gauged, (4) were reviews, (5) were preprints, (6) were book chapters, (7) did not report research on resting-state fcMRI, (8) were not accessible for full text, (9) reported research on data acquired with a field strength under 3.0 T, (10) were theses or dissertations, (11) were meta-analyses, (12) reported research conducted on non-human data, (13) were citation records, (14) were abstract almanacs or miscellaneous publications, (15) were conference papers, (16) were study protocols or methodology papers or (17) were not in English.

Eligibility assessment phase consisted in elimination of articles that did not report metrics that satisfy the criteria of relationality and countability outlined in the Introduction. Eligibility assessment relied on an in-depth inspection of the “Methods” section and a deeper examination of other paper sections in cases where it was necessary. Edge cases were resolved by reviewer consensus.

### 2.5 Study Analysis

#### 2.5.1 Analysis Methods

The sources which passed screening and selection were fully studied. Subsequently, we extracted the metric computation methods reported by the respective authors, described them, and formalized them. To explore the degree of their refinement, state of validation, and level of applicability in a clinical setting, we chose to follow the citations of the articles in question (for better narration consistency and text legibility, these searches will be reported within the Results section). Subsequently, we integrated these findings to yield our statements. We additionally assigned to every metric a Technology Readiness Level (TRL) as specified by ISO 16290:2013 [53] (ISO Standard) in the edition of EU Commission Decision C(2014)4995 [54], better elucidated by us for fcMRI-based abnormality detection applications, on which we will elaborate below. Moreover, we conducted an estimation of computational complexity of the reported methods and used these estimates to perform a complexity-based ranking of the distinct methods. For brevity, within this publication these aspects are reported concisely; detailed information on the complexity estimates and factor magnitudes may be found within the Supplementary Materials.

#### 2.5.2 TRL Assignment Logic

Originally proposed for space systems and published initially as a 7-category scale in 1989 [55], the Technology Readiness Level (TRL) system can be understood as an efficient approach to informing research, development and management decisions for high-precision, high-complexity and/or high-risk systems across various disciplines. By capturing technological maturity according to its ultimate applicability (in our context, clinical diagnostic and prognostic utility), the TRL system thus emerged as an ideal framework for comparing distinct approaches that frequently operate within their own computational and neurobiological paradigms. Here we propose the definitions of the nine levels of technology readiness for fcMRI-based abnormality detection technologies. A tabular subsummation can be found in Table 1.

**Table 1.**
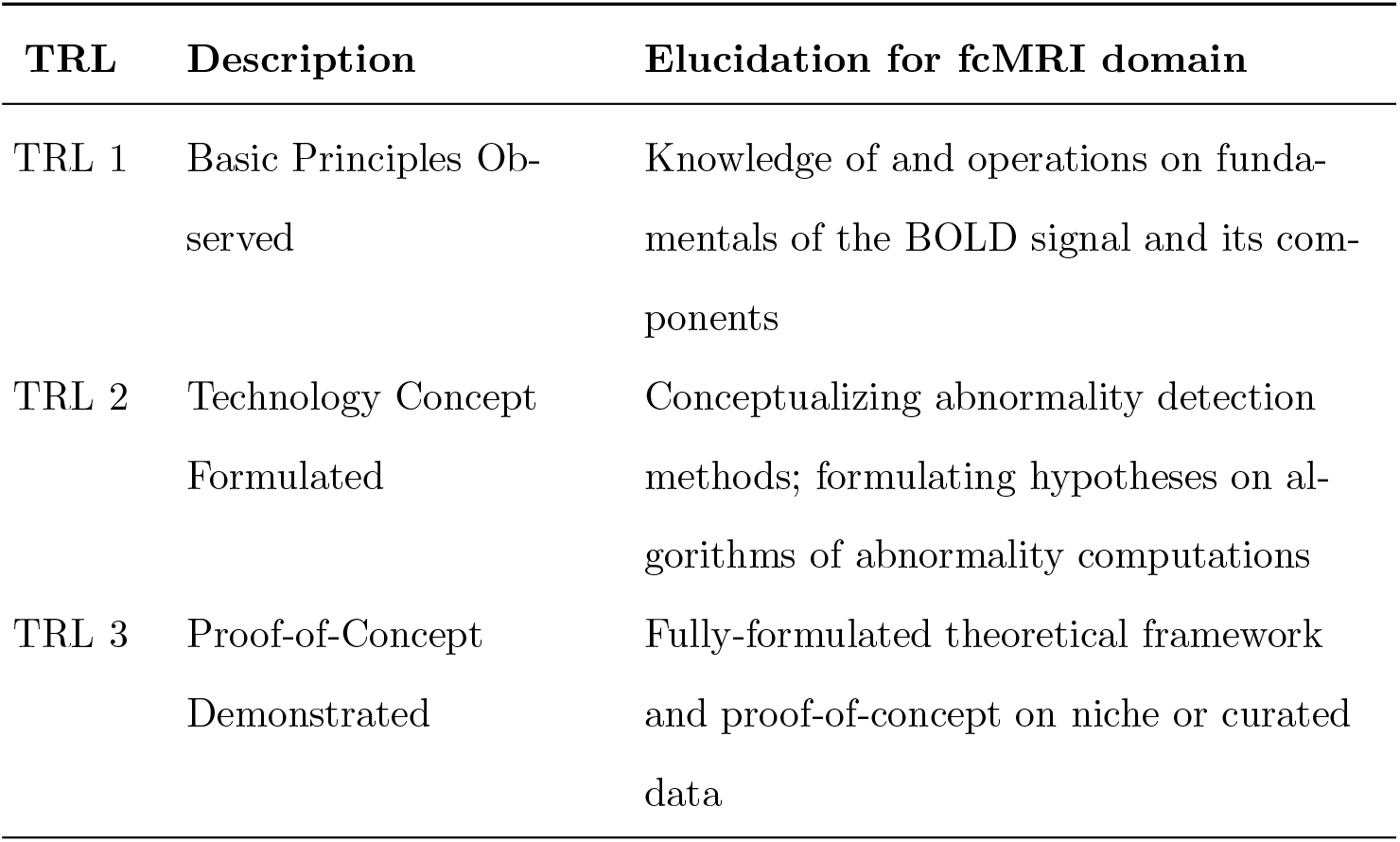
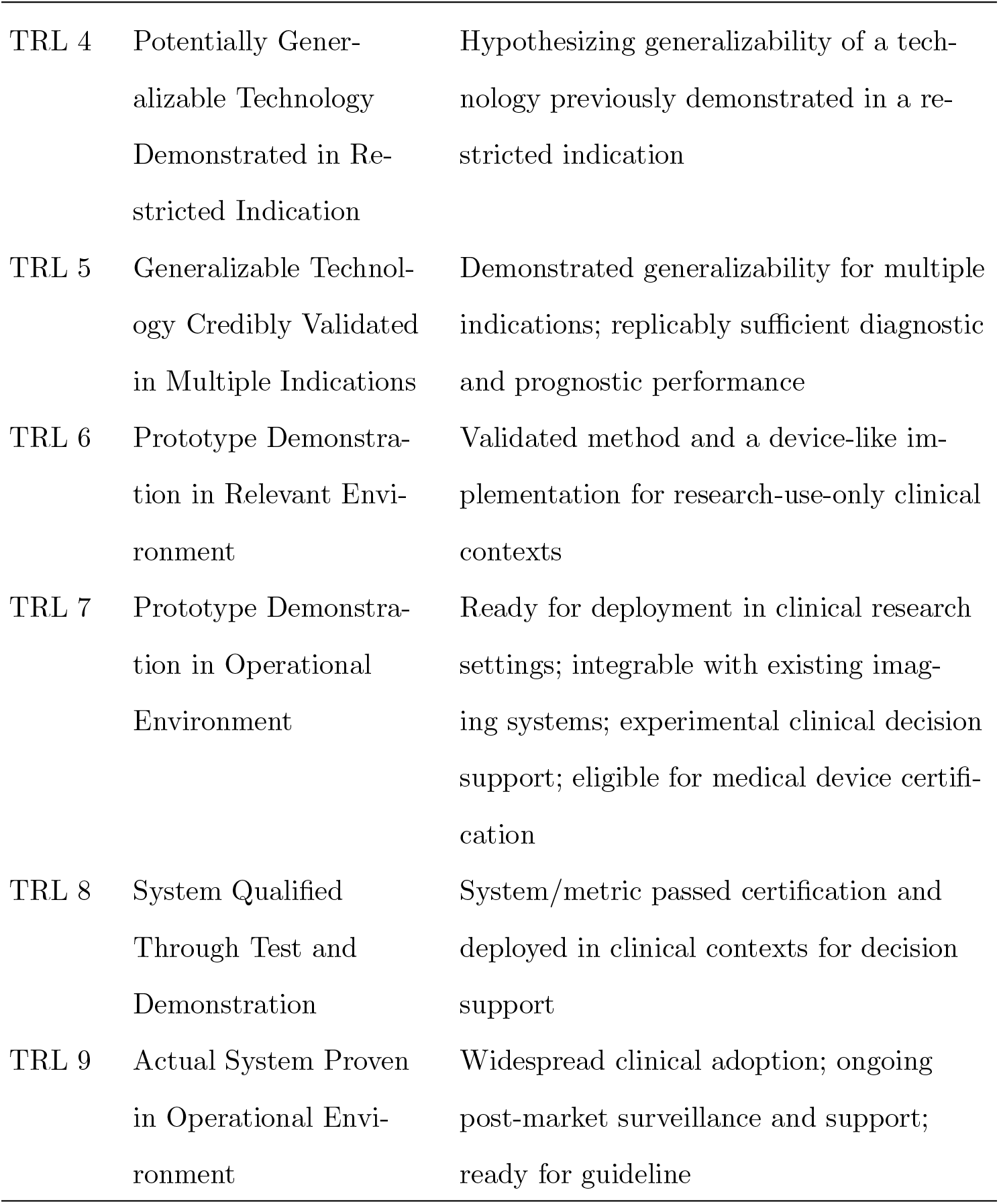
Technology Readiness Levels (TRL) for fcMRI-based abnormality detection.

| TRL | Description | Elucidation for fcMRI domain |
| --- | --- | --- |
| TRL 1 | Basic Principles Ob-<br>served | Knowledge of and operations on funda-<br>mentals of the BOLD signal and its com-<br>ponents |
| TRL 2 | Technology Concept<br>Formulated | Conceptualizing abnormality detection<br>methods; formulating hypotheses on al-<br>gorithms of abnormality computations |
| TRL 3 | Proof-of-Concept<br>Demonstrated | Fully-formulated theoretical framework<br>and proof-of-concept on niche or curated<br>data |
| TRL 4 | Potentially Generalizable Technology Demonstrated in Restricted Indication | Hypothesizing generalizability of a technology previously demonstrated in a restricted indication |
| TRL 5 | Generalizable Technology Credibly Validated in Multiple Indications | Demonstrated generalizability for multiple indications; replicably sufficient diagnostic and prognostic performance |
| TRL 6 | Prototype Demonstration in Relevant Environment | Validated method and a device-like implementation for research-use-only clinical contexts |
| TRL 7 | Prototype Demonstration in Operational Environment | Ready for deployment in clinical research settings; integrable with existing imaging systems; experimental clinical decision support; eligible for medical device certification |
| TRL 8 | System Qualified Through Test and Demonstration | System/metric passed certification and deployed in clinical contexts for decision support |
| TRL 9 | Actual System Proven in Operational Environment | Widespread clinical adoption; ongoing post-market surveillance and support; ready for guideline |

A hypothetical fcMRI abnormality detection technology of level 1 readiness would be operating exclusively with the fundamentals of the BOLD signal, its components and operations regarding their measurement, as well as computation on its direct derivatives.

A level 2 readiness of a hypothetical fcMRI abnormality detection technology implies extensive understanding of the computational and neurobiological basics of fcMRI and may be associated with theoretical proposals of abnormality computations or speculative indications of usability of some parameters for detection of clinical abnormality. Although not easily assignable (as it implies largely theoretical conceptualization and tentative ideation as the technology’s defining element, which is rarely the case for biomedical questions and the publications that accompany them), this readiness level may be exemplified by biomarker search and feature selection studies with a focus on hypothesizing on usability of particular computational measures for diagnostic purposes.

A level 3 of an abnormality detection technology readiness in fcMRI may be defined as the first level with a fully-formulated theoretical framework and proof-of-concept on data pertaining to a niche case or indication without clear understanding of technology’s capacity to be transferred or generalized.

A level 4 of an abnormality detection technology readiness in fcMRI implies generalizability of a technology previously demonstrated in a restricted indication; however, it indicates no such capacity explicitly, requiring further studies in same imaging modality and different clinical indication to explicitly confirm its usability beyond its initial domain.

A level 5 of an abnormality detection technology readiness in fcMRI implies well-demonstrated generalizability for multiple indications and replicably sufficient diagnostic and prognostic performance for potential use in clinical abnormality detection, validated through longitudinal studies. In contrast to the earlier four readiness levels, levels 5 and above imply a scientifically fully-fledged method; however, a level 5 technology does not require a solidified implementation.

Subsequently, a level 6 of an abnormality detection technology readiness in fcMRI may be defined as a first level with a solidified implementation. A level 6 technology is constituted by a validated method and a reported device-like implementation ready for research-use-only (RUO) clinical contexts.

Level 7 of an abnormality detection technology readiness in fcMRI may be defined as the first level that may be structurally and functionally conceptualized as a medical diagnostic device. In contrast to earlier levels, a level 7 technology is fully integrable into the clinical workflow and eligible for medical device certification.

Readiness level 8 implies status post certification and in routine clinical use, while the final level 9 indicates positive results of post-market surveillance.

#### 2.5.3 Computational Complexity Estimation

The computational efficiency of all approaches was estimated as their time (i.e. execution time growth as function of input growth) and space (memory usage growth as function of input growth) complexities. Accurate performance computations and comparisons of algorithms require standardized hardware and data and are determined by implementation language and extent and type of optimization techniques implemented, thus limiting the degree of certainty for formula-based estimates. This circumstance makes it more representative and feasible to hypothesize, estimating the upper bound of asymptotic behavior of algorithms in question by operating within the Big O notation [56]. This approach permits conceptualization of computational pipelines as a sum of individual steps and allows us to approximately gauge complexity of the entire pipeline by appraising the complexity of its most compute-intensive step, making use of the trivial property of the Big O notation:

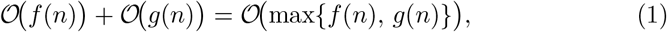

where 𝒪 *f* (*n*) or 𝒪 *g*(*n*) is the set of all functions *f* (*n*) or *g*(*n*) that grow no faster than a constant multiple of some hypothetical functions *h*(*n*) or *m*(*n*) and max(*f* (*n*), *g*(*n*)) is the maximum of the sum of the functions from two sets.

We have based our rankings on the assumption that complexity-defining factors relate to each other in magnitude as

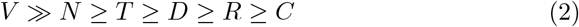

where *V* - number of voxels; *N* - number of individuals; *T* - number of time-points; *D* - number of connectivity features; *R* - number of ROIs/modules/”nodes”; *C* - number of relevant independent components.

It follows that an increment of growth of larger factors would contribute greater to the complexities influenced by these factors, and that products of larger factors or exponents thereof would result in a greater increase of complexities than products or exponents of smaller factors.

#### 2.5.4 Query Keyword Influence Exploration

To gain insight into potential influence of keyword choice on the outcome of our search strategy, we conducted an exploratory re-query in April 2025. We initially input the same keywords and set the query year range to the years 2014-2024, using the Google Scholar result number estimate to obtain a baseline for the number of retrieved publications. We subsequently performed a series of leave-one-out (LOO) queries for each of our keywords, taking note of the result number estimates of the re-query results and the presence of originally located publications. We then stratified the queries along the result number estimates to obtain a representation of each keyword’s contribution to the number of retrieved publications.

## 3 Results

### 3.1 Query Results

Our query cumulatively returned 5696 entries, 5672 of them valid (non-empty, not damaged or fragmentary) entries. After screening, 4964 sources were excluded (Figure 1), while 708 sources were deemed eligible for selection. Only 5 passed selection and were subjected to a full-depth analysis. A PRISMA flow diagram is available in Figure 2.

**Figure 1.**
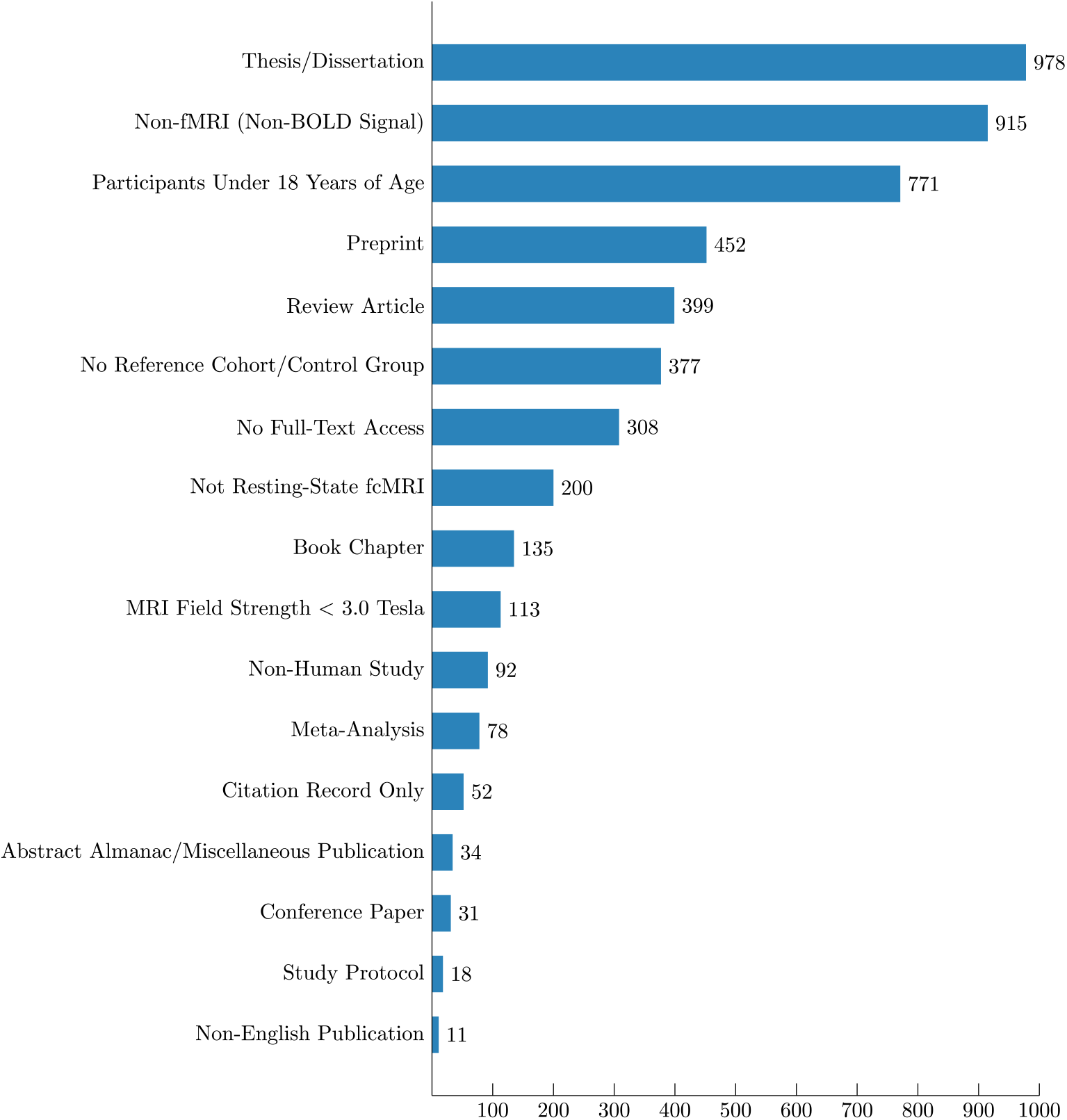
Entries eliminated during Screening phase. In total, we have excluded 4964 entries, of them entries on 978 theses and dissertations, 915 non-fMRI studies, 771 studies on patients under 18 years of age, 452 preprints, 399 reviews, 377 studies without a healthy reference cohort, 308 articles without accessible full-text, 200 non-resting-state fMRI studies, 135 book chapters, 113 studies conducted on data acquired with a field strength under 3.0 Tesla, 92 studies conducted on non-human data, 78 meta-analyses, 52 citation records, 34 abstract almanacs or other publications, 31 conference papers, 18 protocol papers and 11 publications in a language other than English.

**Figure 2.**
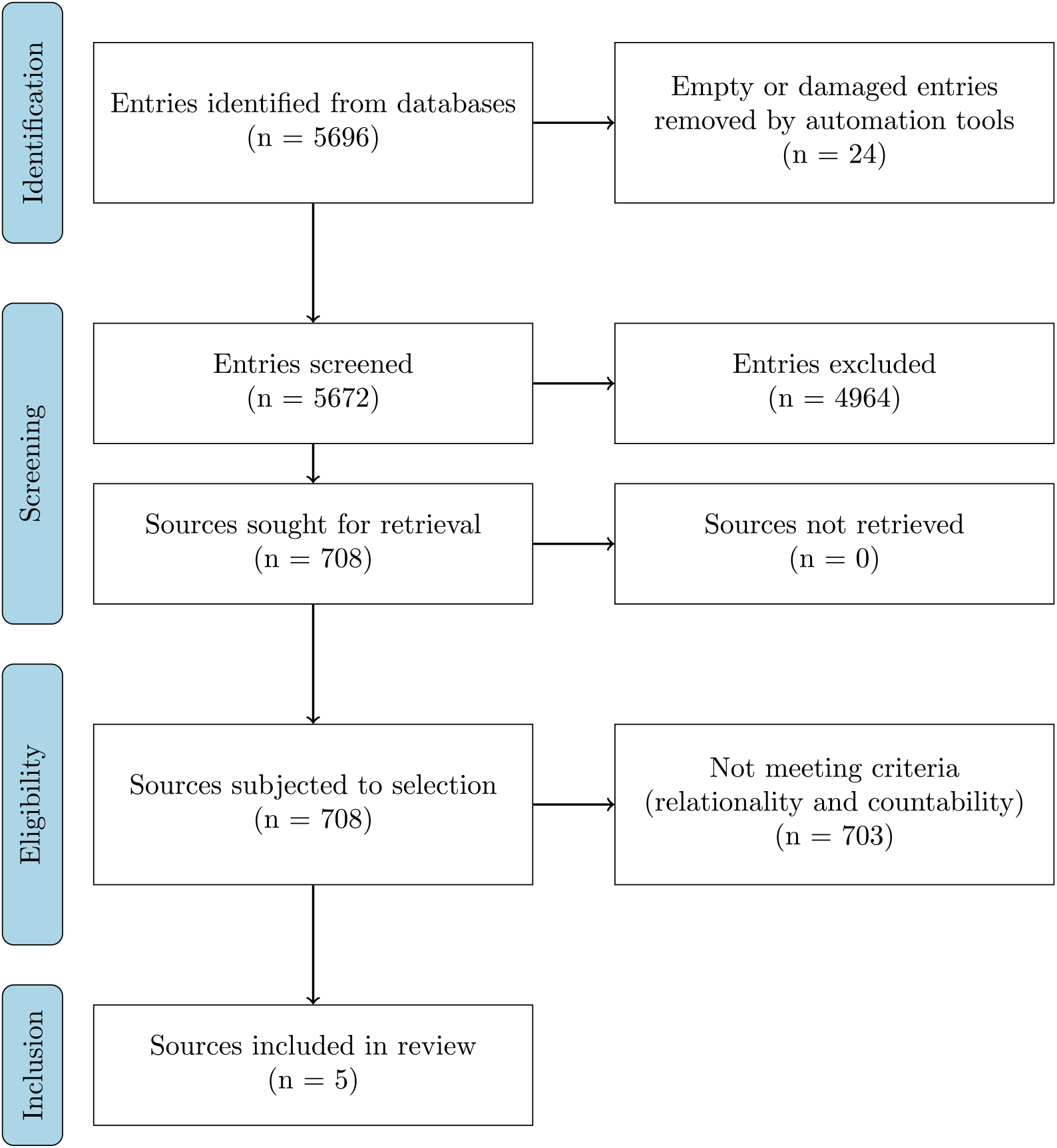
PRISMA flow diagram of review process. In the screening phase we have eliminated 4964 entries of sources, retrieved 708 sources for review eligibility assessment and applied to them the criteria of relationality and countability outlined previously. Notably, only five sources could be deemed eligible for inclusion into the review.

### 3.2 Query Keyword Influence

We have used the number of retrieved publications to obtain a representation of each keyword’s contribution to the total number of retrieved publications (Tab. 2).

**Table 2.**
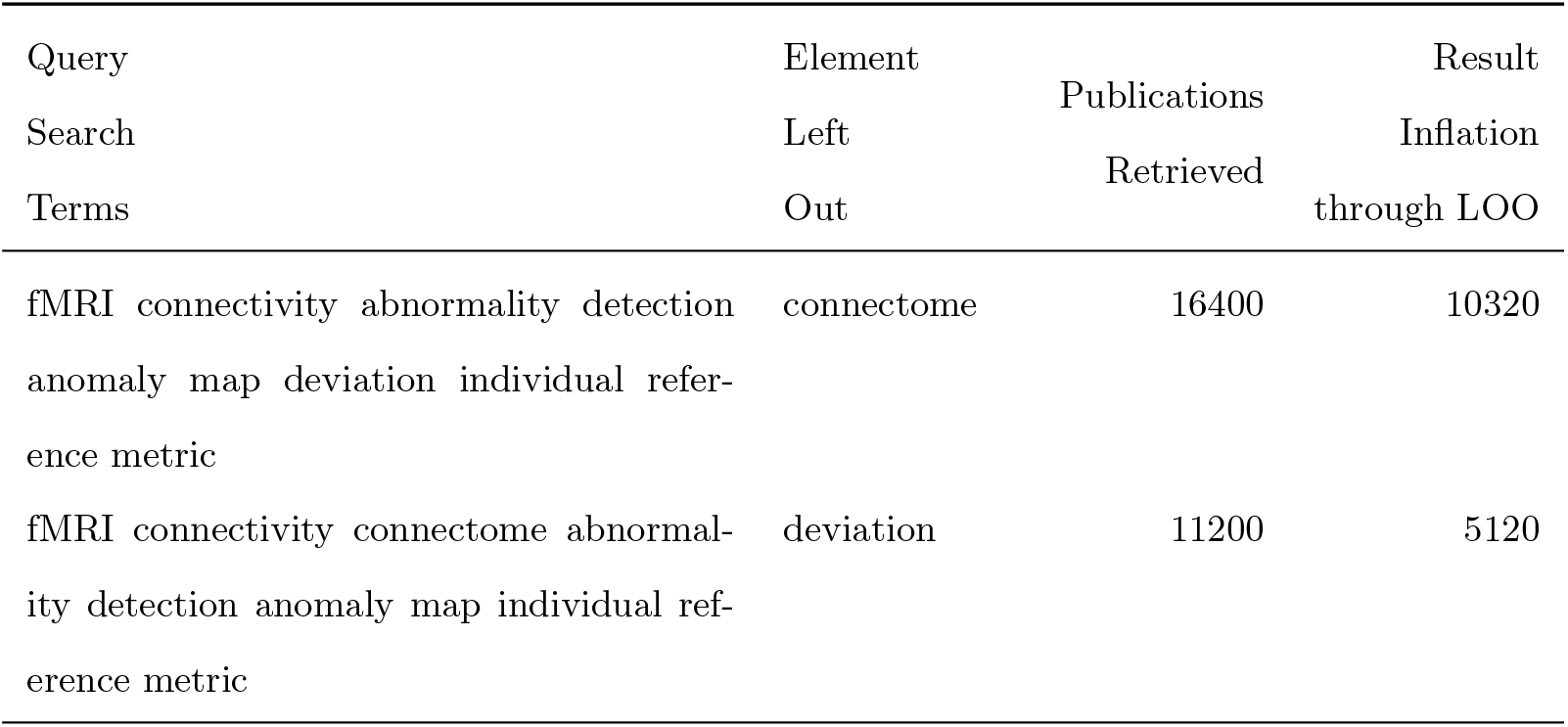
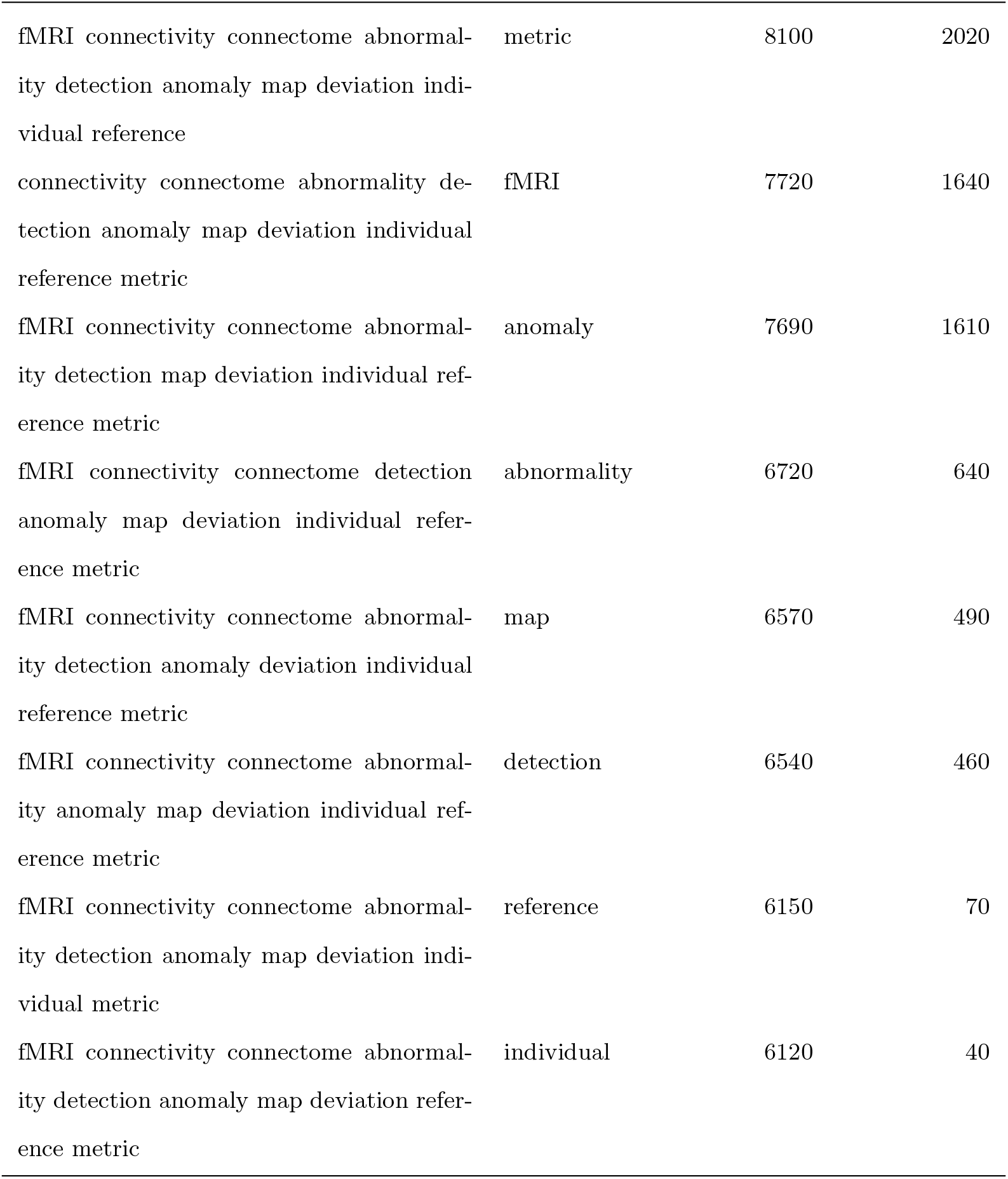
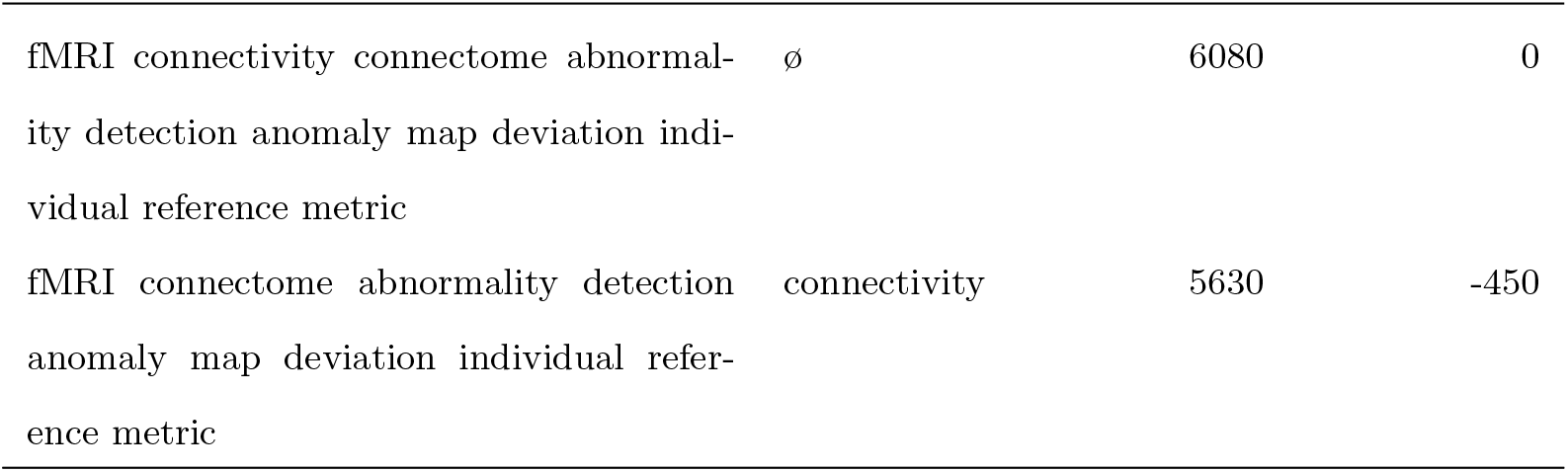
A comparison of query number estimates after a leave-one-out (LOO) query session.

The removal of any but one keyword did not lead to loss of relevant publications we previously reported, but increased the number or retrieved publications, with “connectome”, “deviation”, “metric” and “fMRI” being most responsible for query result inflation. Particularly, just the removal of “connectome” tripled the number of papers returned. Only the removal of “connectivity” deflated the query results.

### 3.3 State of the Art and its Aspects

#### 3.3.1 The Nenning Index

Nenning et al. [57] introduce a voxel-level connectivity abnormality metric in their 2020 glioblastoma paper. Briefly, it has been computed as follows: (1) voxel-wise connectivity matrices for both patients and controls (80 control subjects) are built using z-scored Pearson correlations; (2) element-wise average of control population connectivity matrices is computed to yield a group average “normal” connectivity matrix; (3) a vector of voxel-wise differences is computed between the patients and group average as row-wise cosine similarity; (4) for every voxel in controls’ connectivity matrices and the group average matrix, cosine similarities have been computed to yield voxel-wise distribution; from that distribution, the median and mean absolute deviation (MAD) have been computed (the “voxel mean” and “voxel MAD” respectively); (5) for every patient and for every patient voxel’s cosine similarity, an abnormality score is computed as the difference of cosine similarity and voxel mean, subsequently divided by the voxel MAD.

Analytically, this can be summarized as follows:

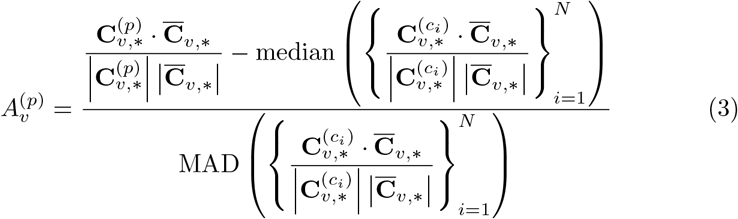

where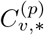is the connectivity vector of voxel *v* for patient *p*,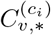is the connectivity vector of voxel *v* for control subject *c*_*i*_, with *i* = 1, 2, …, *N* and *N* = 80 being the number of control subjects,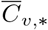 is the average connectivity vector of voxel *v* across all control subjects, ∥· ∥ denotes the Euclidean norm of a vector, · represents the dot product between two vectors, median(·) computes the median of a set of values and MAD(·) computes the median absolute deviation of a set of values.

It is important to mention that Nenning’s team focused on reporting abnormality in non-infiltrated regions but pointed out that the inclusion of tumor infiltrated regions did not significantly alter the overall connectivity signature. Additionally, they demonstrate that in glioblastoma, functional proximity to the tumor tends to be reflected stronger than structural proximity in coefficients derived from fcMRI signal, while visual, somatomotor, and limbic networks tend to exhibit anomaly coefficients more evenly informed by both spatial and functional distance alike. Finally, Nenning’s team demonstrated precedence of network anomalies before tumor recurrence, highlighting a potential diagnostic capacity for abnormality index computation.

PubMed citation check revealed no further studies employing this index in their computations. As the findings of Nenning et al. imply potential generalizability of their index without further validation in other indications, their metric may be assigned a TRL of 4 out of 9.

#### 3.3.2 The Dysconnectivity Index

Stoecklein and Liu [58] present another voxel-level connectivity abnormality metric in their publication on gliomas. It is computed as follows: (1) voxel-wise connectivity matrices are built for both patients and controls (1000 control subjects) using Pearson correlations; (2) for every control subject connectivity matrix, every voxel position in the matrix, and every element in the voxel, a distribution of connectivity coefficients is built; (3) the distribution’s mean and standard deviation are computed to yield respective elements of the mean and standard deviation vectors; (4) for every patient connectivity matrix, every voxel position in the matrix, and every element in the voxel, a z-score is computed for using the elements of the mean and standard deviation vectors computed before (i.e., for i-th element in the patient’s voxel, respective i-th element of the mean and standard deviation vector is used) to yield a vector of z-scores; (5) a sum of z-scores higher than a specific threshold is computed to yield the voxel-level “abnormality coefficient.”

Analytically, for the voxel at the position this can be summarized as follows:

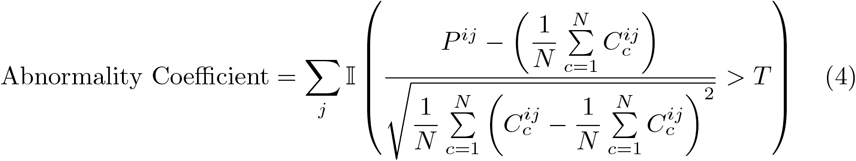

where *P* ^*ij*^ is the connectivity coefficient at voxel position *i, j* for the patient,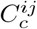 is the connectivity coefficient at voxel position *i, j* for control subject *c, N* is the number of control subjects, *T* is the specific threshold, and 𝕀 (·) is the indicator function, which evaluates to 1 if the condition is true and 0 otherwise.

The authors have conducted computations for the entire brain (without tumor mask exclusion) and demonstrated not only that tumor sites can be captured by their index, but that abnormality can be detected far beyond the lesion itself, even in the contralateral hemisphere, particularly in high grade gliomas. They have also shown that, in glioma, their abnormality index correlates with neurocognitive performance, WHO grade, PET metabolic data, and IDH mutation status. Additionally, the authors hypothesized that abnormal connectivity may not only originate from tumor functional or structural proximity but also indicate sub-clinical tumor cell infiltration and speculated that functional disruption also indicates possible tumor cell infiltration.

PubMed citation check revealed two studies based on this index conducted by either of the two principal researchers. In the first publication [19], the authors demonstrated that their abnormality index (in more recent sources referred to as DCI - the “dysconnectivity index”) can be employed to assess immune effector cell-associated neurotoxicity syndrome (ICANS) in patients under CAR-T therapy and hypothesized that it may be used to objectify damage to functional networks in encephalopathies; furthermore, the authors stated that their index may provide an imaging correlate to trace and possibly predict neurotoxic side- effects of oncologic treatment. In the second publication [59], the authors show a direct association between the DCI and the perifocal edema volume in meningiomas, as well as neurocognitive performance (i.e., higher DCI implies larger edema and more degraded cognition).

The presence of a fully-fledged and scientifically validated method, as well as evidence of index generalizability without further reports of device-like implementation of the Dysconnectivity index allows us to assign to this index a TRL of 5 out of 9.

#### 3.3.3 The Doucet Normative Person-Based Similarity Index

In their publication, Doucet et al. [60] report the normative person-based similarity index (nPBSI). Computed from both functional connectivity and cortical morphometry per aspect, their index explicitly seeks to make a patient’s condition relative to a set control population (93 control subjects). Doucet’s group presents four indices for which clinical, genetic, demographic, and environmental correlates have been described - normative cortical thickness PBSI (nPBSI-CT), normative subcortical volume PBSI (nPBSI-SV), normative module cohesion PBSI (nPBSI-MC) and normative module integrations (nPBSI-MI).

Within the scope of this review, our attention was focused on the BOLD-based module cohesion and module integration metrics, computed as follows: (1) the patient’s brain is parcellated into default mode, central executive, salience, sensorimotor, and visual networks; (2) within-module connectivity is represented as the average value of a voxel wise z-transformed Pearson correlation coefficient between all of the module’s voxel pairs and used to build a patient’s module cohesion profile, encoded as a module cohesion feature vector; (3) between-module connectivity is represented as z-transformed Pearson correlation coefficients of the modules’ averaged time series and used to build a patient’s module integrations profile, encoded as a module integrations feature vector, and finally, (4) the nPBSI-MC or nPBSI-MI are computed as averaged Spearman correlations between the patient and the healthy controls’ respective (module cohesion or module integrations) feature vectors. Analytically, for the patient p this can be summarized as follows:

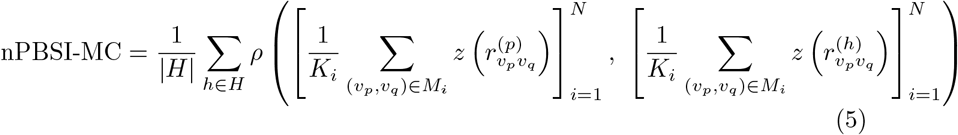

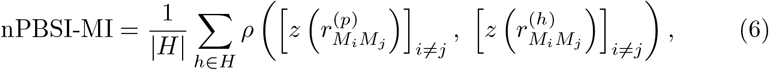

where *N* represents the number of brain modules (default mode, central executive, salience, sensorimotor, and visual networks), *M*_*i*_ is the set of voxels in module *i, K* is the number of voxel pairs in module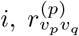 is the Pearson correlation coefficient between voxels *v*_*p*_ and *v*_*q*_ for the patient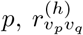 is the Pearson correlation coefficient between voxels *v*_*p*_ and *v*_*q*_ for a healthy control 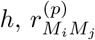 is the Pearson correlation coefficient between the average time series of modules *i* and *j* for the patient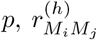is the same for a healthy control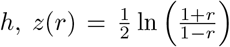is the Fisher z-transformation, *ρ* denotes the Spearman correlation coefficient, *H* is the set of healthy controls and |*H*| is the number of healthy controls.

PubMed citation check revealed no studies employing the normative index from this publication in their computations of functional connectivity metrics. The closest possible match [61] relied on computing both the within and between-network connectivity but did not compute the nPBSI itself. Modest validation for bipolar disorder and lack of nPBSI validation for other disorders constitutes proposal of a theoretical framework with validation in a single indication without implications of generalizability. This circumstance justifies the assignment to this metric of a TRL 3 out of 9.

#### 3.3.4 The Network Topography Spatial Similarity Index

Silvestri and Corbetta present a spatial similarity index (SSI) for network topographies derived from independent component analysis (ICA) in their 2022 publication on gliomas [62]. Briefly, it is computed as follows: (1) rs-fcMRI data of the control population (308 individuals) are subjected to a group ICA (G-ICA) to yield group-level template independent component (IC) maps for ten functional networks (specifically, visual, sensorimotor, auditory, cinguloopercular, dorsal attention, fronto-parietal, default mode, cognitive control, frontal and language networks); (2) the group-level template IC maps are used as spatial constraints for group information-guided ICA (GIG-ICA) of both controls and patients (24 individuals) to produce individual-specific, single-subject level IC maps; (3) for each IC in subject, a cosine similarity is computed between a single-subject IC map and a template IC map thresholded at a value of 1 and yielded as the network topography spatial similarity index.

Analytically, this can be expressed as follows:

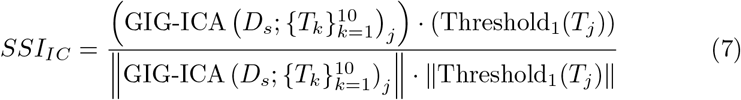

where *SSI*_*IC*_ is the spatial similarity index for a given independent component,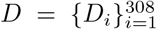 represents the rs-fMRI data of the control population, 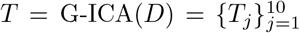are the group-level template IC maps for the ten functional networks obtained from group ICA, *D*_*s*_ is the rs-fMRI data of subject *s*, 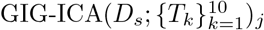produces the single-subject IC map *S*_*s,j*_ for subject *s* and component *j* using the group-level templates as spatial constraints, Threshold_1_(*T*_*j*_) denotes the template IC map *T*_*j*_ thresholded at a value of 1, the numerator (·) represents the dot product between the two vectors and the denominator (∥ · ∥) represents the Euclidean norm (magnitude) of the vectors. The team around Silvestri and Corbetta reported testing structural and functional proximity of their index to the tumor sites, describing partial overlap of index abnormalities and glioma-infiltrated areas and highlighting index abnormalities in non-infiltrated areas. They also analyzed changes in network topography scores and neuropsychological performance and were able to capture a statistically relevant relationship between the SSI and the attention domain.

PubMed citation check revealed no studies employing this normative index in their computations of functional connectivity metrics. Modest validation for gliomas and lack of validation for other disorders constitutes proposal of a theoretical framework with validation in a single indication without implications of generalizability. This circumstance justifies the assignment to this metric of a TRL 3 out of 9.

#### 3.3.5 The Morgan Network Topology Method

Morgan et al. present various metrics and indices in their publication on the role of anterior hippocampus in mesial temporal lobe epilepsy (mTLE) [63]. Their computations rely on multi-modal data and operate within four topologies: the streamline length (*T*_*LEN*_), structural connectivity (*T*_*SC*_), functional connectivity (*T*_*FC*_) and resting-state network topology (*T*_*RSN*_). Within the scope of our review, we will only focus ourselves on the functional connectivity topology and its respective distance index, as no similar index has been reported for the resting-state network topology.

Briefly, it is computed as follows: (1) functional connectivity maps are built for controls (70 individuals) and patients (40 individuals, of them 29 with right mTLE and 11 with left mTLE) from z-transformed functional connectivity matrices through age regression and subsequent averaging of signal over 109 anatomical ROIs; (2) a topology is built from the functional connectivity maps by selecting 55 ROIs of a single hemisphere for patients and controls; (3) a seed vector is used to slice anterior hippocampal connectivity from the topology into a connectivity vector for both patients and controls; (4) the connectivity vector is stratified along connectivity intensity into “bins” to yield their respective connectivity vectors of k elements for both patients and controls; (5) for patient and bin, the Mahalanobis distance between the patient’s bin connectivity vector and the mean of controls’ bin connectivity vectors is computed and yielded as connectivity deviation metric.

Analytically, this can be summarized as follows:

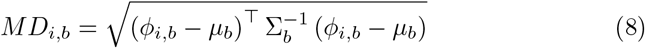

with patient’s connectivity vector in bin, controls’ mean vector in bin and controls’ covariance matrix in bin as, respectively,

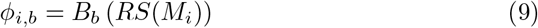

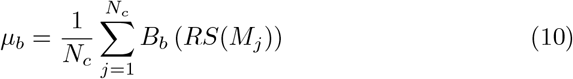

and

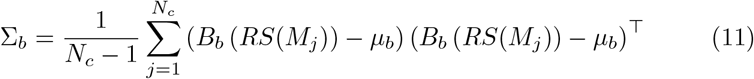

where *M*_*i*_ is a functional connectivity matrix (size 109 × 109) for individual *i, S*(*M*_*i*_) denotes a selection operator extracting a 55 × 55 hemisphere-specific submatrix from *M*_*i*_, *R* is a seed vector (size 1 × 55) with 1 at the anterior hippocampus position and 0 elsewhere, *B*_*b*_(·) symbolizes the binning function that selects elements belonging to bin *b* based on connectivity intensity, *N*_*c*_ = 70 is the number of control individuals, *µ*_*b*_ is the mean vector of controls’ connectivity vectors in bin *b* and Σ_*b*_ is the covariance matrix of controls’ connectivity vectors in bin *b*.

PubMed citation check revealed two studies which reported intriguing use of the logic behind this computational approach. The first publication of interest by Morgan et al. [64] reports use of similar connectivity profiling techniques and the Mahalanobis distance for outcome prediction in mTLE patents by means of distance computation between a patient’s connectivity profile and a normative population of individuals who achieved seizure-free status after mesial temporal lobe surgery. Notably, the team around Morgan reported sensitivity of 100% and specificity of 90% for their prediction approach.

The second publication by Guerrero-Gonzalez et al. [65] does not pertain to functional MRI, but uses the comparable logic of normative modeling and Mahalanobis distance to quantify abnormality in tractography of traumatic brain injury patients.

The epilepsy-specific focus of Morgan’s distance-based approach limits the scope of potential use of this metric; however, success of similar computational approaches in other modalities and remarkable performance of the Mahalanobis distance-based index in the surgical outcome prediction task allows us to contstate the Morgan’s method as a theoretical framework with validation that implies generalizability potential without supplying stronger evidence. This supports the assignment to this metric of a TRL 4 out of 9.

### 3.4 Computational Complexity Estimation

We have found that voxel-wise metrics and their naïve computational methods, such as the DCI, are associated with the highest computational cost, while methods strongly relying on a-priori hypotheses for dimensionality reduction, such as the NTM, are the least costly. This phenomenon was particularly prominent for the time complexity, which tended to grow with decreasing levels of functional and anatomical abstraction. Simultaneously, in all but naïve methods the bulk of space complexity was constituted by the static element (i.e. initially present data).

At minimal abstraction levels (i.e. at voxel-wise LOD), both time and space complexities increased exponentially to the voxel resolution of input data. With the voxel count of a typical fcMRI image tensor greatly outscaling any other factors (also see 2.5.3), it follows intuitively that any voxel-based or voxel-derived (i.e. sufficiently granular ROI-based) computing methods are inherently and immediately associated with steeper system requirements.

A ranking of the metric computation methods can be found in (Fig. 3).

**Figure 3.**
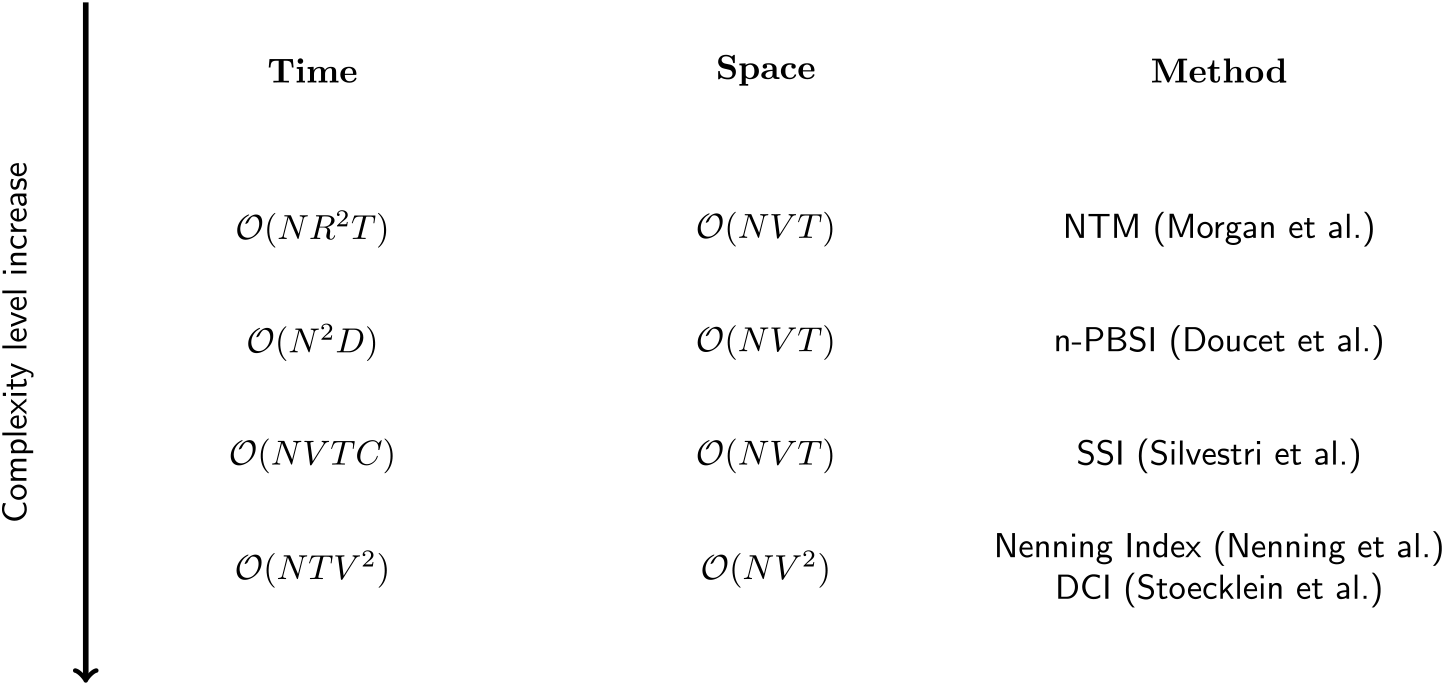
Ranking of reported methods by their estimated time and space complexities. Lower position indicates higher time/space complexity and increased projected computational cost growth as a function of input. Denominations: N - number of individuals; D - number of connectivity features; R - number of ROIs; T - number of timepoints; V - number of voxels; C - number of relevant independent components; n-PBSI - normative Person-Based Similarity Index [60]); NTM - network topology method [63]; SSI - spatial similarity index [62] (Silvestri et al., 2022); DCI - dysconnectivity index [58].

## 4 Discussion

### 4.1 Group Comparison Currently Prevails in Studies of Abnormal Connectivity

In this review, we have been able to show that, despite the strong knowledge base to support the concept of neurodegenerative [8, 9], psychiatric [10] and neuro-oncological diseases [11, 12, 13] as “network disorders”, a metric capable of evaluating and quantifying large-scale functional brain network disruptions in individual patients is yet to be developed, validated and made accessible enough for potential incorporation into clinical practice.

We also demonstrated that, despite the significant benefits of relational metrics as integral elements of normative modeling [66], only five such metrics of functional connectivity deviation have been proposed within the last ten years. Of note, in many studies that we evaluated for this review, the findings and the hypotheses that lead to these findings were built around the aspiration to illustrate a binary difference between patients and healthy controls, which resulted in reports of metrics being increased or decreased in patients, often without a clearly specified relation between the increment of metric and increment of pathological state.

The development of patient-centric fcMRI markers requires moving beyond group comparison and toward relational metrics based on normative populations that span variability in demographic and procedural factors.

### 4.2 Specifics, Limitations and Use Cases of Current Abnormality Quantification Metrics

#### 4.2.1 Lesion Data Handling

The five reported indices are variably informed by signals from lesional tissue. To this end, a distinction between lesion-informed, lesion-uninformed and lesionagnostic indices may be proposed. Below, we will define the three types by illustrating their features as exemplified by the reported metrics.

An example of a lesion-uninformed index is constituted by the Nenning index [57]. Not informed by the data from the macroscopically-visible glioma-infiltrated regions, this index was sought by the authors to quantify abnormal functional connectivity in regions distant from the main tumor mass. As also mentioned by the authors themselves, the principal merit of such approach consists in greater sensitivity to actual discordant connectivity, as the index would remain unperturbed by the tumor-associated loss of axonal connections and the respective signal changes.

As opposed to the lesion-uninformed type, the concept of a lesion-informed index is exemplified by the dysconnectivity index [58]), the network topography spatial similarity index [62] and the Morgan network topology method [63]. As the name suggests, the lesion signal (tumor tissue signal, as in the former two cases, or hippocampal sclerosis signal, as in the latter case) was included into computations and directly influenced the indices presented. The two most obvious and principal merits of a lesion-informed index would consist in the reduced modularity of its computational pipeline and greater practicability for its clinical context due to its inherent capacity to quantify connectivity abnormalities both within and outside the lesion.

Finally, the lesion-agnostic class of index is embodied by the normative person-based similarity index [60] (Doucet et al., 2020), developed specifically to capture abnormal connectivity in bipolar disorder and excluding lesional data. Its ability to capture and quantify connectivity abnormality in conditions associated with structural lesions remains unclear, thus justifying its categorization into a class of its own.

#### 4.2.2 ROI Level of Detail

The five indices at hand operate within various levels of detail defined both by their clinical indications and their computational approaches. For instance, the voxel-wise indices accept and yield data on finer-grained voxels that continuously span the entire gray matter, while the non-voxel-wise indices make use of preconceptions to achieve various degrees of abstraction.

The dysconnectivity index [58]), and the Nenning index [57] operate at the voxel level. Both indices employ voxel-wise computations to yield voxel-wise metrics of abnormal connectivity and, consequently, allow for higher granularity of results, potentially lending themselves well to diagnosis as well as planning and control of therapy. Moreover, these indices can inform calculation of larger-scale measures with a capacity to inform more specialized experimental approaches.

Other indices exhibit various levels of inherent granularity, using their apriori hypotheses and assumptions for dimensionality reduction so as to quantify abnormal connectivity against a normative population at a lower computational cost. For instance, the Morgan network topology method [63] operates on a scale of ROIs (also dubbed “nodes” in a later publication [64]), which are determined by the Multi-Atlas algorithm [67, 68] and FreeSurfer [69], and is slightly less granular than the voxel-wise indices. Similarly, the network topography spatial similarity index [62], operating on independent component analysis, makes use of Yeo-7 [70] and Gordon [71] atlases to determine component affiliation with a particular functional network. Lastly, the normative person-based similarity index [60] in its module integrations and module cohesion variation employs as sources for time-course averaging the “modules” taken from regions as per [72, 73] to yield its whole-brain measure.

#### 4.2.3 Synthesis: Potential Clinical Use Cases for Indices and Combinations Thereof

Drawing upon our previous definitions, findings and observations, we can delineate distinct use cases for the index classes presented above and propose an approach to combining them in routine diagnostic practice.

Herein, the lesion-informed, voxel-wise-LOD naïve (“general”) indices may be of highest utility in clinical diagnostic contexts, when the necessity of exploring the connectomic underpinnings of a clinical syndrome clearly justifies the significant computational cost. Moreover, they may be more generalizable due to the minimal amount of abstraction within their core logic (i.e. minimal dependence on an a-priori clinical hypothesis), thus potentially lending themselves best to validation across many disorders. Finally, as they can be readily abstracted into coarser derivatives, they may be able to inform clinical decision-making more objectively.

In contrast, other (“specialist”) index classes may be used most optimally for special indications or for longitudinal quantification of previously-diagnosed conditions, as their higher initial abstraction implies the necessity of distinct apriori clinical hypotheses, which may limit generalizability and validation across multiple disorders, but positively influences computational complexity and practical repeatability.

Computing both index classes implies indication for data acquisition in the form of a clinical syndrome with a set of associated a-priori hypotheses and differential diagnoses. Hence, a tandem of a “general” metric and one or more “specialist” metrics could be both diagnostically beneficial and logistically feasible, constituting a case of maximized hardware utilization. This can be substantiated by the fact that the “general” time/space complexity outscales the “specialist” time/space complexities exponentially (see Results), thus defining the system requirements. Consequently, “specialist” indices could be attached to sufficiently optimized “general” pipelines at a fraction of “general” time complexity without significant inflation of space complexity and with an increase in clinical utility through production of multi-metric index-signatures.

In summary, the clinical utility for the patient may be feasibly maximized through building of optimized multi-metric diagnostic pipelines with one invariant “general” (naïve) voxel-wise metric and variant “specialist” (custom-LOD) metrics determined by the patient’s condition to produce multi-metric index-signatures.

### 4.3 Strategies for Improvement of Current and Emerging Abnormality Quantification Metrics

#### 4.3.1 Methodical Strategies for Improvement of Current and Emerging Abnormality Quantification Methods

The current circumstances create a uniquely favorable setting for more practical progress on relational fcMRI-based metrics of abnormal connectivity, making possible the implementation of a set of strategies aimed to develop the SOTA in fcMRI-based abnormal connectivity quantification, which we will propose below.

Firstly, a considerable acceleration of progress in fcMRI-based metric research can be achieved through creation of large-scale reference datasets augmented by technical and demographic parameters. Currently, high-quality data can be accessed freely by virtue of recognized cohorts [39, 40, 41] and openaccess data repositories [42], which permits compilation of harmonized, statistically powerful reference datasets, capturing variability across demographics and technical parameters. As the utility of accounting for these factors is well-substantiated by evidence of variables such as age [74, 75, 76], sex [77] and scan parameters [78, 25] having significant influence on fcMRI metrics, having better access to data on these factors would permit greater insight into biological and technical variability and help inform more refined and nuanced normative models with potentially greater utility for clinical application.

Moreover, the utility of each present and new metric can be better established through high-quality validation (i.e. study designs accommodating for confident statements on metric utility) in their respective conditions. Herein, longitudinal analyses with a focus on determining relation between the increment of metric and the increment of pathological state are most desirable. In neuro-oncological and neurodegenerative conditions a study of the abnormality metric dynamic with a focus on areas exhibiting morphological abnormality or those within structural or functional proximity of such areas (as in e.g. [57]) with simultaneous examination of associations with PET, neurocognitive testing, clinical endpoints such as overall survival (as in [18]) and genetic profiling of tissue (as in e.g. [58]) would constitute an optimal strategy. Indices in psychiatric conditions may be best validated by focusing on relation between metric increments and symptom severity score increments, treatment monitoring or prediction of treatment response. In chemical-, toxinor infection-related conditions the relation between metric and pathological state increments would call for a mode of analysis similar to such in neuro-oncological conditions, with the key parameters measured against metric increments being determined by the condition in question, e.g. ICE (Immune Effector Cell Encephalopathy), and EEG abnormalities (as in e.g. [19]).

Finally, better reproducibility and comparability of current and emerging methods may be achieved through adoption of benchmark datasets for performance measurements and greater disclosure of the algorithms and their code implementation to peer review. This would permit improved collaboration and assist in creation of a consolidated methodological paradigm, fostering consensus on potential clinical indications and the fcMRI-derived metrics best suited to address them.

#### 4.3.2 Computational Strategies for Improvement of Current and Emerging Abnormality Quantification Methods

The advent of big data and artificial intelligence-based methods in fcMRI research may boost the development of relational connectivity metrics by enhancing the current computational approaches.

The drastic progress in computing technology [79] has made possible the widespread use of industrial-grade hardware acceleration of previously strictly linear computing through parallel computing with the help of much more readily accessible graphical processing units (GPUs) [81, 80]. Improved hardware-software synergy now permits optimization of both speed and efficiency of data engineering and machine learning, allowing for faster simultaneous read/write operations and deeper insight into highly complex multidimensional data. This is well-manifested by the packages for accelerated Python computing (e.g. CuPy and Dask [82, 83]), optimized tensor storage solutions (e.g. Zarr or Xarray [84, 85]), new Neuroimaging Informatics Technology Initiative (NifTI) image manipulation modules (e.g. Xibabel [86]) or the advancements in the field of machine learning (ML) frameworks [87, 89, 88] and hyper-parallel computing (e.g. the Bend programming language, based on HVM2 [90, 91]).

Moreover, the current rise of deep learning models for operations on fcMRI data can help streamline previously time-consuming elements of data preprocessing and enrichment, potentially accelerating research on relational fcMRI-based metrics manyfold. This is prominently exemplified by ML breakthroughs in the area of structural image preprocessing with algorithms such as FastSurfer [92], a deep learning pipeline for brain segmentation, cortical surface reconstruction, cortical label mapping and thickness analyses. Similar advancements have also been reported for affine registration with tools such as SynthMorph [93], a model that resolves a tensor-to-tensor mapping problem for an image pair, yielding a compatible spatial transform. Lastly, experimental ML-boosted integrated pipelines for fcMRI image preprocessing (e.g. DeepPrep [94]) have also been reported.

Lastly, methods of patient- and condition-specific denoising strategies with a potential to improve the overall signal quality of fcMRI data are currently emerging and may be incorporated into future preprocessing pipelines [35].

Employing the aforementioned measures to optimize computational complexity and maximize algorithm performance in current and emerging fcMRI-based metric computing pipelines may help increase feasibility of their use in clinical contexts.

### 4.4 Study Limitations

#### 4.4.1 Query Time Range

Our search only comprises sources released before mid-May 2024.

#### 4.4.2 Neurobiology-Related Constraints

Due to considerably less generalizable dynamics of neurobiological development in pediatric and adolescent individuals, a decision was made not to consider publications that concerned persons under 18 years of age.

#### 4.4.3 Search Terms, Peer Review Constraints, and Keyword Influence

Our search terms might not include all relevant publications. In particular, preprints, theses and dissertations have been excluded as reports that have not undergone a peer review process.

Another factor constituting a source of limitations can be our choice of keywords. As shown in Results, the removal of all but one individual keyword did not lead to loss of originally located publications, but led to decreased query specificity, inflating the number of retrieved papers by up to three times per removed keyword. This illustrates that the least restrictive query does not necessarily yield optimal results, as also discussed in [95]. Simultaneously, upon removal of “connectivity”, the retrieval number decreased, indicating a less sensitive retrieval and implicating potential loss of relevant publications. Taken together, this simulation may substantiate our choice of keywords.

#### 4.4.4 Exclusion of Publications Based on Task fcMRI Data

Even though task-based fcMRI may capture more behaviorally relevant information [96] and was reported to exhibit superior information gain in comparison to rs-fcMRI, particularly for the Theory-of-Mind task [97], we decided to focus on metrics based on resting-state fcMRI (rs-fcMRI) for this review, as we deem these indices more suitable for routine clinical diagnostics than those based on task fcMRI (t-fcMRI) for several reasons. Firstly, t-fcMRI’s overall test-retest reliability remains variable, depending on the respective task paradigm [98, 99, 100]. Secondly, task-related networks can also be assessed using rs-fcMRI as task-related states may be conceptualized as derivatives of intrinsic network activity, as substantiated by evidence of intrinsic networks being highly similar to resting-state networks in architecture and consistent across task paradigms in their activity [101]. Moreover, task-based fcMRI acquisition requires from the patient sufficient language proficiency, cognitive ability, and compliance, which drastically limits potential clinical applicability of t-fcMRI and its derivative metrics. Finally, t-fcMRI possesses a set of special logistical requirements, most notably MRI-compatible audiovisual equipment and accompanying technical expertise in use and maintenance, both of which are predominantly available only in specialized centers. These circumstances thus substantiate choice of rs-fcMRI-derived over t-fcMRI-derived metrics as feasible for wide application in routine diagnostics.

## 5 Summary

Patients suffering from neuro-oncological, psychiatric and neurodegenerative disorders can benefit from individualized detection and quantification of abnormal functional connectivity. However, no fcMRI-derived imaging markers have yet seen widespread adoption in clinical research or practice. Within the scope of this review, we have asserted both the necessity and the current absence of a well-established relational and countable metric for abnormal functional connectivity in individuals. We have subsequently leveraged the Google Scholar database to retrieve sources that matched our search criteria and subjected them to PRISMA-compliant screening and selection to yield items for subsequent indepth analysis. We then demonstrated five currently reported methods/metrics for relational, normative quantification of abnormal connectivity, formalized their computation methods, assessed their Technology Readiness Level and estimated their computational efficiency. Building upon our findings, we have discussed the metrics’ lesion data handling, region of interest level of detail and the potential clinical use cases. We reaffirmed the need of moving beyond group comparison and toward quantitative fcMRI anomaly metrics for application in individual patients and proposed methodical and computational strategies for improvement of current and emerging abnormality quantification metrics.

## Acknowledgements

We would like to thank Julia Ruat for her invaluable support in the management of this project.

## Data Availability Statement

Data sharing is not applicable in the context of this publication, as no datasets were generated or analyzed during this scoping review. The tabular reports of the included and excluded articles are available from the corresponding authors upon reasonable request.

## Code Availability Statement

No novel code was generated during the current study. Minimal scripting was done to support data aggregation.

## Inclusion and Ethics Statement

This scoping review concerns peer-reviewed publications and therefore does not require ethical approval.

## Author Contributions

A. T. - Conceptualization, Methodology Selection & Implementation, Data Collection, Entry Screening, Source Eligibility Selection, Source Analysis, Formalization & Integration of Findings, Original Draft Preparation, Visualization, Review and Editing, Project Administration.

D. V. - Data Collection, Entry Screening, Source Eligibility Selection, Source Analysis, Formalization & Integration of Findings, Original Draft Preparation, Visualization, Review and Editing, Project Administration.

H. K. - Data Collection, Entry Screening, Source Eligibility Selection, Source Analysis, Formalization & Integration of Findings, Original Draft Preparation, Visualization, Review and Editing, Project Administration.

R. L. - Entry Screening, Source Eligibility Selection, Source Analysis, Formalization & Integration of Findings.

P. M. - Entry Screening, Source Eligibility Selection.

D. R. - Entry Screening, Source Eligibility Selection.

A. D. - Entry Screening.

A. V. - Entry Screening.

V. P. - Entry Screening.

S. W. - Source Eligibility Selection, Source Analysis.

S. S. - Conceptualization, Source Analysis, Formalization & Integration of Findings, Review and Editing, Supervision, Funding Acquisition, Project Ad- ministration, Resources, Oversight and Approvals.

D.V. and H.K. contributed equally to this publication.

D.R. and P.M. contributed equally to this publication.

A.D. and A.V. contributed equally to this publication.

## References

Biswal, B., Yetkin, F. Z., Haughton, V. M., & Hyde, J. S. (1995). Functional connectivity in the motor cortex of resting human brain using echo-planar mri. Magnetic Resonance in Medicine, 34(4), 537–541. 10.1002/mrm.1910340409

Ogawa, S., Lee, T. M., Kay, A. R., & Tank, D. W. (1990). Brain magnetic resonance imaging with contrast dependent on blood oxygenation. Proceedings of the National Academy of Sciences, 87(24), 9868–9872. 10.1073/pnas.87.24.9868

Logothetis, N. K. (2003). The underpinnings of the BOLD Functional Magnetic Resonance Imaging signal. Journal of Neuroscience, 23(10), 3963–3971. 10.1523/jneurosci.23-10-03963.2003

Buxton, R. B., Wong, E. C., & Frank, L. R. (1998). Dynamics of blood flow and oxygenation changes during brain activation: The balloon model. Magnetic Resonance in Medicine, 39(6), 855–864. 10.1002/mrm.1910390602

Buckner, R. L., Krienen, F. M., & Yeo, B. T. T. (2013). Opportunities and limitations of intrinsic functional connectivity MRI. Nature Neuroscience, 16(7), 832–837. 10.1038/nn.3423

Zhang, J., Kucyi, A., Raya, J., Nielsen, A. N., Nomi, J. S., Damoiseaux, J. S., Greene, D. J., Horovitz, S. G., Uddin, L. Q., & Whitfield-Gabrieli, S. (2021). What have we really learned from functional connectivity in clinical populations? NeuroImage, 242, 118466. 10.1016/j.neuroimage.2021.118466

Pagani, M., Gutierrez-Barragan, D., De Guzman, A. E., Xu, T., & Gozzi, A. (2023). Mapping and comparing fMRI connectivity networks across species. Communications Biology, 6(1). 10.1038/s42003-023-05629-w

Franzmeier, N., Dewenter, A., Frontzkowski, L., Dichgans, M., Rubinski, A., Neitzel, J., Smith, R., Strandberg, O., Ossenkoppele, R., Buerger, K., Duering, M., Hansson, O., & Ewers, M. (2020). Patient-centered connectivitybased prediction of tau pathology spread in Alzheimer’s disease. Science Advances, 6(48). 10.1126/sciadv.abd1327

Rauchmann, B., Brendel, M., Franzmeier, N., Trappmann, L., Zaganjori, M., Ersoezlue, E., Morenas-Rodriguez, E., Guersel, S., Burow, L., Kurz, C., Haeckert, J., Tató, M., Utecht, J., Papazov, B., Pogarell, O., Janowitz, D., Buerger, K., Ewers, M., Palleis, C., … Perneczky, R. (2022). Microglial activation and connectivity in Alzheimer disease and aging. Annals of Neurology, 92(5), 768–781. 10.1002/ana.26465

Georgiadis, F., Lariviére, S., Glahn, D., Hong, L. E., Kochunov, P., Mowry, B., Loughland, C., Pantelis, C., Henskens, F. A., Green, M. J., Cairns, M. J., Michie, P. T., Rasser, P. E., Catts, S., Tooney, P., Scott, R. J., Schall, U., Carr, V., Quidé, Y., … Kirschner, M. (2024). Connectome architecture shapes large-scale cortical alterations in schizophrenia: a worldwide ENIGMA study. Molecular Psychiatry, 29(6), 1869–1881. 10.1038/s41380-024-02442-7

Winkler, F., Venkatesh, H. S., Amit, M., Batchelor, T., Demir, I. E., Deneen, B., Gutmann, D. H., Hervey-Jumper, S., Kuner, T., Mabbott, D., Platten, M., Rolls, A., Sloan, E. K., Wang, T. C., Wick, W., Venkataramani, V., & Monje, M. (2023). Cancer neuroscience: State of the field, emerging directions. Cell, 186(8), 1689–1707. 10.1016/j.cell.2023.02.002

Hausmann, D., Hoffmann, D. C., Venkataramani, V., Jung, E., Horschitz, S., Tetzlaff, S. K., Jabali, A., Hai, L., Kessler, T., Azórin, D. D., Weil, S., Kourtesakis, A., Sievers, P., Habel, A., Breckwoldt, M. O., Karreman, M. A., Ratliff, M., Messmer, J. M., Yang, Y., … Winkler, F. (2022). Autonomous rhythmic activity in glioma networks drives brain tumour growth. Nature, 613(7942), 179–186. 10.1038/s41586-022-05520-4

Salvalaggio, A., Pini, L., Bertoldo, A., & Corbetta, M. (2024). Glioblastoma and brain connectivity: the need for a paradigm shift. The Lancet Neurology, 23(7), 740–748. 10.1016/s1474-4422(24)00160-1

Fox, M. D., Liu, H., & Pascual-Leone, A. (2012). Identification of reproducible individualized targets for treatment of depression with TMS based on intrinsic connectivity. NeuroImage, 66, 151–160. 10.1016/j.neuroimage.2012.10.082

Brys, M., Fox, M. D., Agarwal, S., Biagioni, M., Dacpano, G., Kumar, P., Pirraglia, E., Chen, R., Wu, A., Fernandez, H., Shukla, A. W., Lou, J., Gray, Z., Simon, D. K., Di Rocco, A., & Pascual-Leone, A. (2016). Multifocal repetitive TMS for motor and mood symptoms of Parkinson disease. Neurology, 87(18), 1907–1915. 10.1212/wnl.0000000000003279

Ren, J., Ren, W., Zhou, Y., Dahmani, L., Duan, X., Fu, X., Wang, Y., Pan, R., Zhao, J., Zhang, P., Wang, B., Yu, W., Chen, Z., Zhang, X., Sun, J., Ding, M., Huang, J., Xu, L., Li, S., … Liu, H. (2023). Personalized functional imaging-guided rTMS on the superior frontal gyrus for post-stroke aphasia: A randomized sham-controlled trial. Brain Stimulation, 16(5), 1313–1321. 10.1016/j.brs.2023.08.023

Cui, W., Wang, Y., Ren, J., Hubbard, C. S., Fu, X., Fang, S., Wang, D., Zhang, H., Li, Y., Li, L., Jiang, T., & Liu, H. (2022). Personalized fMRI Delineates Functional Regions Preserved within Brain Tumors. Annals of Neurology, 91(3), 353–366. 10.1002/ana.26303

Sprugnoli, G., Rigolo, L., Faria, M., Juvekar, P., Tie, Y., Rossi, S., Sverzellati, N., Golby, A. J., & Santarnecchi, E. (2022). Tumor BOLD connectivity profile correlates with glioma patients’ survival. Neuro-Oncology Advances, 4(1). 10.1093/noajnl/vdac153

Stoecklein, S., Wunderlich, S., Papazov, B., Winkelmann, M., Kunz, W. G., Mueller, K., Ernst, K., Stoecklein, V. M., Blumenberg, V., Karschnia, P., Bücklein, V. L., Rejeski, K., Schmidt, C., Von Bergwelt-Baildon, M., Tonn, J., Ricke, J., Liu, H., Remi, J., Subklewe, M., … Schoeberl, F. (2023). Functional connectivity MRI provides an imaging correlate for chimeric antigen receptor T-cell-associated neurotoxicity. Neuro-Oncology Advances, 5(1). 10.1093/noajnl/vdad135

Sjuls, G. S., & Specht, K. (2022). Variability in Resting-State functional magnetic resonance imaging: the effect of body mass, blood pressure, hematocrit, and glycated hemoglobin on hemodynamic and neuronal parameters. Brain Connectivity, 12(10), 870–882. 10.1089/brain.2021.0125

Duncan, N., & Northoff, G. (2013). Overview of potential procedural and participantrelated confounds for neuroimaging of the resting state. Journal of Psychiatry and Neuroscience, 38(2), 84–96. 10.1503/jpn.120059

Brennan, D., Murrough, J. W., & Morris, L. S. (2020). Intrasubject functional connectivity related to self-generated thoughts. Brain and Behavior, 11(1). 10.1002/brb3.1860

Han, X., Jovicich, J., Salat, D., Van Der Kouwe, A., Quinn, B., Czanner, S., Busa, E., Pacheco, J., Albert, M., Killiany, R., Maguire, P., Rosas, D., Makris, N., Dale, A., Dickerson, B., & Fischl, B. (2006). Reliability of MRI-derived measurements of human cerebral cortical thickness: The effects of field strength, scanner upgrade and manufacturer. NeuroImage, 32(1), 180–194. 10.1016/j.neuroimage.2006.02.051

Jovicich, J., Czanner, S., Han, X., Salat, D., Van Der Kouwe, A., Quinn, B., Pacheco, J., Albert, M., Killiany, R., & Blacker, D. (2009). MRI-derived measurements of human subcortical, ventricular and intracranial brain volumes: Reliability effects of scan sessions, acquisition sequences, data analyses, scanner upgrade, scanner vendors and field strengths. NeuroImage, 46(1), 177–192. 10.1016/j.neuroimage.2009.02.010

Mueller, S., Wang, D., Fox, M. D., Pan, R., Lu, J., Li, K., Sun, W., Buckner, R. L., & Liu, H. (2015). Reliability correction for functional connectivity: Theory and implementation. Human Brain Mapping, 36(11), 4664–4680. 10.1002/hbm.22947

Reeder, S. B., Atalar, E., Bolster, B. D., & McVeigh, E. R. (1997). Quantification and reduction of ghosting artifacts in interleaved echo-planar imaging. Magnetic Resonance in Medicine, 38(3), 429–439. 10.1002/mrm.1910380312

Takao, H., Hayashi, N., & Ohtomo, K. (2011). Effect of scanner in longitudinal studies of brain volume changes. Journal of Magnetic Resonance Imaging, 34(2), 438–444. 10.1002/jmri.22636

Triantafyllou, C., Hoge, R., Krueger, G., Wiggins, C., Potthast, A., Wiggins, G., & Wald, L. (2005). Comparison of physiological noise at 1.5 T, 3 T and 7 T and optimization of fMRI acquisition parameters. NeuroImage, 26(1), 243–250. 10.1016/j.neuroimage.2005.01.007

Triantafyllou, C., Polimeni, J. R., & Wald, L. L. (2010). Physiological noise and signal-to-noise ratio in fMRI with multi-channel array coils. NeuroImage, 55(2), 597–606. 10.1016/j.neuroimage.2010.11.084

Kayvanrad, A., Arnott, S. R., Churchill, N., Hassel, S., Chemparathy, A., Dong, F., Zamyadi, M., Gee, T., Bartha, R., Black, S. E., Lawrence-Dewar, J. M., Scott, C. J., Symons, S., Davis, A. D., Hall, G. B., Harris, J., Lobaugh, N. J., MacQueen, G., Woo, C., & Strother, S. (2021). Resting state fMRI scanner instabilities revealed by longitudinal phantom scans in a multi-center study. NeuroImage, 237, 118197. 10.1016/j.neuroimage.2021.118197

Teeuw, J., Pol, H. E. H., Boomsma, D. I., & Brouwer, R. M. (2021). Reliability modelling of resting-state functional connectivity. NeuroImage, 231, 117842. 10.1016/j.neuroimage.2021.117842

Zhang, C., Baum, S. A., Adduru, V. R., Biswal, B. B., & Michael, A. M. (2018). Test-retest reliability of dynamic functional connectivity in resting state fMRI. NeuroImage, 183, 907–918. 10.1016/j.neuroimage.2018.08.021

Van Dijk, K. R., Sabuncu, M. R., & Buckner, R. L. (2011). The influence of head motion on intrinsic functional connectivity MRI. NeuroImage, 59(1), 431–438. 10.1016/j.neuroimage.2011.07.044

Siegel, J. S., Mitra, A., Laumann, T. O., Seitzman, B. A., Raichle, M., Corbetta, M., & Snyder, A. Z. (2016). Data quality influences observed links between functional connectivity and behavior. Cerebral Cortex, 27(9), 4492–4502. 10.1093/cercor/bhw253

Wunderlich, S., Alici, C., Motevallia, S., Stoecklein, V., Von Baumgarten, L., Schoeberl, F., Subklewe, M., Schulz, E., & Stoecklein, S. (2025). Denoising Strategies of Functional Connectivity MRI data in Lesional and Non-Lesional Brain Diseases. medRxiv (Cold Spring Harbor Laboratory). 10.1101/2025.01.22.25320407

Lindquist, M. A., Geuter, S., Wager, T. D., & Caffo, B. S. (2019). Modular preprocessing pipelines can reintroduce artifacts into fMRI data. Human Brain Mapping, 40(8), 2358–2376. 10.1002/hbm.24528

Luppi, A. I., Gellersen, H. M., Liu, Z., Peattie, A. R. D., Manktelow, A. E., Adapa, R., Owen, A. M., Naci, L., Menon, D. K., Dimitriadis, S. I., & Stamatakis, E. A. (2024). Systematic evaluation of fMRI data-processing pipelines for consistent functional connectomics. Nature Communications, 15(1). 10.1038/s41467-024-48781-5

Cho, J. W., Korchmaros, A., Vogelstein, J. T., Milham, M. P., & Xu, T. (2020). Impact of concatenating fMRI data on reliability for functional connectomics. NeuroImage, 226, 117549. 10.1016/j.neuroimage.2020.117549

David C. Van Essen, Stephen M. Smith, Deanna M. Barch, Timothy E.J. Behrens, Essa Yacoub, Kamil Ugurbil, for the WU-Minn HCP Consortium. (2013). The WU-Minn Human Connectome Project: An overview. NeuroImage 80(2013):62-79.

Petersen, R. C., Aisen, P. S., Beckett, L. A., Donohue, M. C., Gamst, A. C., Harvey, D. J., Jack, C. R., Jagust, W. J., Shaw, L. M., Toga, A. W., Trojanowski, J. Q., & Weiner, M. W. (2009). Alzheimer’s Disease Neuroimaging Initiative (ADNI). Neurology, 74(3), 201–209. 10.1212/wnl.0b013e3181cb3e25

Holmes, A. J., Hollinshead, M. O., O’Keefe, T. M., Petrov, V. I., Fariello, G. R., Wald, L. L., Fischl, B., Rosen, B. R., Mair, R. W., Roffman, J. L., Smoller, J. W., & Buckner, R. L. (2015). Brain Genomics Superstruct Project initial data release with structural, functional, and behavioral measures. Scientific Data, 2(1). 10.1038/sdata.2015.31

OpenNeuro. (10.05.25). https://openneuro.org/

Chamberland, M., Genc, S., Tax, C. M. W., Shastin, D., Koller, K., Raven, E. P., Cunningham, A., Doherty, J., Van Den Bree, M. B. M., Parker, G. D., Hamandi, K., Gray, W. P., & Jones, D. K. (2021). Detecting microstructural deviations in individuals with deep diffusion MRI tractometry. Nature Computational Science, 1(9), 598–606. 10.1038/s43588-02100126-8

Nugent, S., Croteau, E., Potvin, O., Castellano, C., Dieumegarde, L., Cunnane, S. C., & Duchesne, S. (2020). Selection of the optimal intensity normalization region for FDG-PET studies of normal aging and Alzheimer’s disease. Scientific Reports, 10(1). 10.1038/s41598-020-65957-3

López-González, F. J., Silva-Rodríguez, J., Paredes-Pacheco, J., Niñerola-Baizán, A., Efthimiou, N., Martín-Martín, C., Moscoso, A., Ruibal, Á., Roé-Vellvé, N., & Aguiar, P. (2020). Intensity normalization methods in brain FDG-PET quantification. NeuroImage, 222, 117229. 10.1016/j.neuroimage.2020.117229

Arksey, H., & O’Malley, L. (2005). Scoping studies: towards a methodological framework. International Journal of Social Research Methodology, 8(1), 19–32. 10.1080/1364557032000119616

Levac, D., Colquhoun, H., & O’Brien, K. K. (2010). Scoping studies: advancing the methodology. Implementation Science, 5(1). 10.1186/1748-5908-5-69

Tricco, A. C., Lillie, E., Zarin, W., O’Brien, K. K., Colquhoun, H., Levac, D., Moher, D., Peters, M. D., Horsley, T., Weeks, L., Hempel, S., Akl, E. A., Chang, C., McGowan, J., Stewart, L., Hartling, L., Aldcroft, A., Wilson, M. G., Garritty, C., … Straus, S. E. (2018). PRISMA Extension for Scoping Reviews (PRISMA-SCR): Checklist and explanation. Annals of Internal Medicine, 169(7), 467–473. 10.7326/m18-0850

Harzing, A.W. (2007) Publish or Perish, available from https://harzing.com/resources/publish-or-perish

McKinney, W. (2010). Data structures for statistical computing in Python. Proceedings of the Python in Science Conferences, 56–61. 10.25080/majora-92bf1922-00a

Harris, C. R., Millman, K. J., Van Der Walt, S. J., Gommers, R., Virtanen, P., Cournapeau, D., Wieser, E., Taylor, J., Berg, S., Smith, N. J., Kern, R., Picus, M., Hoyer, S., Van Kerkwijk, M. H., Brett, M., Haldane, A., Del Río, J. F., Wiebe, M., Peterson, P., … Oliphant, T. E. (2020). Array programming with NumPy. Nature, 585(7825), 357–362. 10.1038/s41586-020-2649-2

Your connected workspace for wiki, docs & projects — Notion. (10.05.25). Notion. https://www.notion.so/

ISO 16290:2013. (10.05.25). ISO. https://www.iso.org/standard/56064.html

Research and innovation. (10.05.25). European Commission. https://ec.europa.eu/research/participants/data/ref/h2020/other/wp/2018-2020/annexes/h2020-wp1820-annex-g-trl_en.pdf

Sadin, S. R., Povinelli, F. P., & Rosen, R. (1989). The NASA technology push towards future space mission systems. Acta Astronautica, 20, 73–77. 10.1016/0094-5765(89)90054-4

Cormen, T. H., Leiserson, C. E., Rivest, R. L., & Stein, C. (2022b). Introduction to Algorithms, fourth edition. MIT Press.

Nenning, K., Furtner, J., Kiesel, B., Schwartz, E., Roetzer, T., Fortelny, N., Bock, C., Grisold, A., Marko, M., Leutmezer, F., Liu, H., Golland, P., Stoecklein, S., Hainfellner, J. A., Kasprian, G., Prayer, D., Marosi, C., Widhalm, G., Woehrer, A., & Langs, G. (2020). Distributed changes of the functional connectome in patients with glioblastoma. Scientific Reports, 10(1). 10.1038/s41598-020-74726-1

Stoecklein, V. M., Stoecklein, S., Galié, F., Ren, J., Schmutzer, M., Unterrainer, M., Albert, N. L., Kreth, F., Thon, N., Liebig, T., Ertl-Wagner, B., Tonn, J., & Liu, H. (2020). Resting-state fMRI detects alterations in whole brain connectivity related to tumor biology in glioma patients. NeuroOncology, 22(9), 1388–1398. 10.1093/neuonc/noaa044

Stoecklein, V., Wunderlich, S., Papazov, B., Thon, N., Schmutzer, M., Schinner, R., Zimmermann, H., Liebig, T., Ricke, J., Liu, H., Tonn, J., Schichor, C., & Stoecklein, S. (2023). Perifocal Edema in Patients with Meningioma is Associated with Impaired Whole-Brain Connectivity as Detected by Resting-State fMRI. American Journal of Neuroradiology, 44(7), 814–819. 10.3174/ajnr.a7915

Doucet, G. E., Glahn, D. C., & Frangou, S. (2020). Person-based similarity in brain structure and functional connectivity in bipolar disorder. Journal of Affective Disorders, 276, 38–44. 10.1016/j.jad.2020.06.041

West, A., Hamlin, N., Frangou, S., Wilson, T. W., & Doucet, G. E. (2021). Person-Based Similarity Index for Cognition and its neural correlates in Late Adulthood: Implications for Cognitive Reserve. Cerebral Cortex, 32(2), 397–407. 10.1093/cercor/bhab215

Silvestri, E., Moretto, M., Facchini, S., Castellaro, M., Anglani, M., Monai, E., D’Avella, D., Della Puppa, A., Cecchin, D., Bertoldo, A., & Corbetta, M. (2022). Widespread cortical functional disconnection in gliomas: an individual network mapping approach. Brain Communications, 4(2). 10.1093/braincomms/fcac082

Morgan, V. L., Johnson, G. W., Cai, L. Y., Landman, B. A., Schilling, K. G., Englot, D. J., Rogers, B. P., & Chang, C. (2021). MRI network progression in mesial temporal lobe epilepsy related to healthy brain architecture. Network Neuroscience, 5(2), 434–450. 10.1162/netn_a_00184

Morgan, V. L., Sainburg, L. E., Johnson, G. W., Janson, A., Levine, K. K., Rogers, B. P., Chang, C., & Englot, D. J. (2022). Presurgical temporal lobe epilepsy connectome fingerprint for seizure outcome prediction. Brain Communications, 4(3). 10.1093/braincomms/fcac128

Guerrero-Gonzalez, J. M., Yeske, B., Kirk, G. R., Bell, M. J., Ferrazzano, P. A., & Alexander, A. L. (2022). Mahalanobis distance tractometry (MaD-Tract) – a framework for personalized white matter anomaly detection applied to TBI. NeuroImage, 260, 119475. 10.1016/j.neuroimage.2022.119475

Marquand, A. F., Kia, S. M., Zabihi, M., Wolfers, T., Buitelaar, J. K., & Beckmann, C. F. (2019). Conceptualizing mental disorders as deviations from normative functioning. Molecular Psychiatry, 24(10), 1415–1424. 10.1038/s41380-019-0441-1

Asman, A. J., & Landman, B. A. (2012). Non-local statistical label fusion for multi-atlas segmentation. Medical Image Analysis, 17(2), 194–208. 10.1016/j.media.2012.10.002

Huo, Y., Plassard, A. J., Carass, A., Resnick, S. M., Pham, D. L., Prince, J. L., & Landman, B. A. (2016). Consistent cortical reconstruction and multi-atlas brain segmentation. NeuroImage, 138, 197–210. 10.1016/j.neuroimage.2016.05.030

Fischl, B. (2012). FreeSurfer. NeuroImage, 62(2), 774–781. 10.1016/j.neuroimage.2012.01.021

Yeo, B. T. T., Krienen, F. M., Sepulcre, J., Sabuncu, M. R., Lashkari, D., Hollinshead, M., Roffman, J. L., Smoller, J. W., Zöllei, L., Polimeni, J. R., Fischl, B., Liu, H., & Buckner, R. L. (2011). The organization of the human cerebral cortex estimated by intrinsic functional connectivity. Journal of Neurophysiology, 106(3), 1125–1165. 10.1152/jn.00338.2011

Gordon, E. M., Laumann, T. O., Adeyemo, B., Huckins, J. F., Kelley, W. M., & Petersen, S. E. (2014). Generation and Evaluation of a Cortical Area Parcellation from Resting-State Correlations. Cerebral Cortex, 26(1), 288–303. 10.1093/cercor/bhu239

Zalesky, A., Fornito, A., & Bullmore, E. T. (2010). Network-based statistic: Identifying differences in brain networks. NeuroImage, 53(4), 1197–1207. 10.1016/j.neuroimage.2010.06.041

Crossley, N. A., Mechelli, A., Vértes, P. E., Winton-Brown, T. T., Patel, A. X., Ginestet, C. E., McGuire, P., & Bullmore, E. T. (2013). Cognitive relevance of the community structure of the human brain functional coactivation network. Proceedings of the National Academy of Sciences, 110(28), 11583–11588. 10.1073/pnas.1220826110

Farras-Permanyer, L., Mancho-Fora, N., Montalá-Flaquer, M., Bartrés-Faz, D., Vaqué-Alcázar, L., Peró-Cebollero, M., & Guárdia-Olmos, J. (2019). Age-related changes in resting-state functional connectivity in older adults. Neural Regeneration Research, 14(9), 1544. 10.4103/16735374.255976

Geerligs, L., Renken, R. J., Saliasi, E., Maurits, N. M., & Lorist, M. M. (2014). A Brain-Wide study of Age-Related changes in functional connectivity. Cerebral Cortex, 25(7), 1987–1999. 10.1093/cercor/bhu012

Andrews-Hanna, J. R., Snyder, A. Z., Vincent, J. L., Lustig, C., Head, D., Raichle, M. E., & Buckner, R. L. (2007). Disruption of Large-Scale brain systems in advanced aging. Neuron, 56(5), 924–935. 10.1016/j.neuron.2007.10.038

David, S. P., Naudet, F., Laude, J., Radua, J., Fusar-Poli, P., Chu, I., Stefanick, M. L., & Ioannidis, J. P. A. (2018). Potential reporting bias in neuroimaging studies of sex differences. Scientific Reports, 8(1). 10.1038/s41598-018-23976-1

Chen, A. A., Srinivasan, D., Pomponio, R., Fan, Y., Nasrallah, I. M., Resnick, S. M., Beason-Held, L. L., Davatzikos, C., Satterthwaite, T. D., Bassett, D. S., Shinohara, R. T., & Shou, H. (2022). Harmonizing functional connectivity reduces scanner effects in community detection. NeuroImage, 256, 119198. 10.1016/j.neuroimage.2022.119198

Coyle, D., & Hampton, L. (2023). 21st century progress in computing. Telecommunications Policy, 48(1), 102649. 10.1016/j.telpol.2023.102649

AMD Technical Information Portal. (04.12.24). https://docs.amd.com/v/u/en-US/wp505-versal-acap

NVIDIA RTX Series Datasheets. (04.12.24). NVIDIA. https://resources.nvidia.com/en-us-briefcase-for-datasheets/

R. Okuta, Y. Unno, D. Nishino, S. Hido, and C. Loomis. CuPy: A NumPy-Compatible Library for NVIDIA GPU Calculations. Proceedings of Work-shop on Machine Learning Systems (LearningSys) in The Thirty-first Annual Conference on Neural Information Processing Systems (NIPS), 2017 http://learningsys.org/nips17/assets/papers/paper_16.pdf

Dask Development Team (2016). Dask: Library for dynamic task scheduling URL http://dask.pydata.org

Alistair Miles, jakirkham, Joe Hamman, Dimitri Papadopoulos Orfanos, M Bussonnier, Josh Moore, David Stansby, Davis Bennett, Tom Augspurger, James Bourbeau, Andrew Fulton, Sanket Verma, Deepak Cherian, Norman Rzepka, Ryan Abernathey, Gregory Lee, Mads R. B. Kristensen, Zain Patel, Saransh Chopra, … Shivank Chaudhary. (2024). zarr-developers/zarr-python: v3.0.0-beta.2 (v3.0.0-beta.2). Zenodo. 10.5281/zenodo.14165945

Hoyer, S., & Hamman, J. (2017). xarray: N-D labeled Arrays and Datasets in Python. Journal of Open Research Software, 5(1), 10. 10.5334/jors.148

Matthew-Brett. (04.12.24). GitHub -matthew-brett/xibabel: Piloting a new image object for neuroimaging based on XArray. GitHub. https://github.com/matthew-brett/xibabel

Jax-Ml. (04.12.24). GitHub - jax-ml/jax: Composable transformations of Python+NumPy programs: differentiate, vectorize, JIT to GPU/TPU, and more. GitHub. http://github.com/jax-ml/jax

Martín Abadi, Ashish Agarwal, Paul Barham, Eugene Brevdo, Zhifeng Chen, Craig Citro, Greg S. Corrado, Andy Davis, Jeffrey Dean, Matthieu Devin, Sanjay Ghemawat, Ian Goodfellow, Andrew Harp, Geoffrey Irving, Michael Isard, Rafal Jozefowicz, Yangqing Jia, Lukasz Kaiser, Manjunath Kudlur, Josh Levenberg, Dan Mané, Mike Schuster, Rajat Monga, Sherry Moore, Derek Murray, Chris Olah, Jonathon Shlens, Benoit Steiner, Ilya Sutskever, Kunal Talwar, Paul Tucker, Vincent Vanhoucke, Vijay Vasudevan, Fernanda Viégas, Oriol Vinyals, Pete Warden, Martin Wattenberg, Martin Wicke, Yuan Yu, and Xiaoqiang Zheng. TensorFlow: Large-scale machine learning on heterogeneous systems, 2015. https://tensorflow.org.

Paszke, A., Gross, S., Massa, F., Lerer, A., Bradbury, J., Chanan, G., Killeen, T., Lin, Z., Gimelshein, N., Antiga, L., Desmaison, A., Köpf, A., Yang, E., DeVito, Z., Raison, M., Tejani, A., Chilamkurthy, S., Steiner, B., Fang, L., … Chintala, S. (2019). PyTorch: An Imperative Style, High-Performance Deep Learning Library. arXiv (Cornell University). 10.48550/arxiv.1912.01703

HVM2.pdf. (10.05.2025). https://docs.google.com/viewer?url=https://raw.githubusercontent.com/Higher

Higher order company. (10.05.2025). https://higherorderco.com/

Henschel, L., Conjeti, S., Estrada, S., Diers, K., Fischl, B., & Reuter, M. (2020). FastSurfer - A fast and accurate deep learning based neuroimaging pipeline. NeuroImage, 219, 117012. 10.1016/j.neuroimage.2020.117012

Hoffmann, M., Hoopes, A., Greve, D. N., Fischl, B., & Dalca, A. V. (2024). Anatomy-aware and acquisition-agnostic joint registration with SynthMorph. Imaging Neuroscience, 2, 1–33. 10.1162/imag_a_00197

Ren, J., An, N., Lin, C., Zhang, Y., Sun, Z., Zhang, W., Li, S., Guo, N., Cui, W., Hu, Q., Wang, W., Wu, X., Wang, Y., Jiang, T., Satterthwaite, T. D., Wang, D., & Liu, H. (2025). DeepPrep: an accelerated, scalable and robust pipeline for neuroimaging preprocessing empowered by deep learning. Nature Methods. 10.1038/s41592-025-02599-1

Scells, H., Forbes, C., Clark, J., Koopman, B., & Zuccon, G. (2022). The impact of query refinement on systematic review Literature search. ICTIR ‘22: Proceedings of the 2022 ACM SIGIR International Conference on Theory of Information Retrieval. 10.1145/3539813.3545143

Zhao, W., Makowski, C., Hagler, D. J., Garavan, H. P., Thompson, W. K., Greene, D. J., Jernigan, T. L., & Dale, A. M. (2023). Task fMRI paradigms may capture more behaviorally relevant information than resting-state functional connectivity. NeuroImage, 270, 119946. 10.1016/j.neuroimage.2023.119946

Tuominen, J., Specht, K., Vaisvilaite, L., & Zeidman, P. (2023). An information-theoretic analysis of resting-state versus task fMRI. Network Neuroscience, 7(2), 769–786. 10.1162/netna00302

Bennett, C. M., & Miller, M. B. (2013). fMRI reliability: Influences of task and experimental design. Cognitive Affective & Behavioral Neuroscience, 13(4), 690–702. 10.3758/s13415-013-0195-1

Rai, S., Graff, K., Tansey, R., & Bray, S. (2024). How do tasks impact the reliability of fMRI functional connectivity? Human Brain Mapping, 45(3). 10.1002/hbm.26535

Savage, H. S., Mulders, P. C. R., Van Eijndhoven, P. F. P., Van Oort, J., Tendolkar, I., Vrijsen, J. N., Beckmann, C. F., & Marquand, A. F. (2024). Dissecting task-based fMRI activity using normative modelling: an application to the Emotional Face Matching Task. Communications Biology, 7(1). 10.1038/s42003-024-06573-z

Cole, M. W., Bassett, D. S., Power, J. D., Braver, T. S., & Petersen, S. E. (2014). Intrinsic and Task-Evoked network architectures of the human brain. Neuron, 83(1), 238–251. 10.1016/j.neuron.2014.05.014

