## Supplementary Material for "Quantification of Brain Functional Connectivity Deviations in Individuals: A Scoping Review of Functional MRI Studies"

### Supplementary Material: Computational Complexity and Complexity-Defining Factor Magnitude Estimates

May 2025

### 1 Introduction

#### 1.1 General Considerations

Here we will discuss in depth the possible computational complexity of the reported algorithms. To this end, we introduce a set of definitions, assumptions and denominations, use these to estimate the order of magnitude of individual factors that may define the computational complexity, justify our estimations and build upon this framework to establish a theoretical estimate for the computational complexities of the individual algorithms.

#### 1.2 Big O Notation

The Big O notation is a common method in asymptotic analysis used for denoting the upper bound of the function's asymptotic behavior [1]. As it is a "worst case" notation (implying description of the least possibly optimized behavior of a program), it is the type of notation most suitable for our use case.

The Big O notation postulates that a function exhibiting asymptotic behavior grows no faster than its fastest-growing element:

$$\mathcal{O}\big(f(n)\big) + \mathcal{O}\big(g(n)\big) = \mathcal{O}\big(\max\{f(n),\, g(n)\}\big), \tag{1}$$

where $\mathcal{O}\big(f(n)\big)$ or $\mathcal{O}\big(g(n)\big)$ is the set of all functions $f(n)$ or $g(n)$ that grow no faster than a constant multiple of some hypothetical functions $h(n)$ or $m(n)$ and $\max(f(n), g(n))$ is the maximum of the sum of the functions from two sets.

This property allows us to focus ourselves on the most complex elements of the algorithms reported.

#### 1.3 Factors Defining Computational Complexity

The extent of a method's complexities and aspects of its tentative clinical applicability are determined by the numerical magnitude of its individual contributing factors. During our analysis of the five metrics, we have observed that the complexities may be most influenced by voxel amount, number of individual subjects (particularly, number of individuals included into the normative sample), amount of time points within the fcMRI data, amount of ROIs involved in the ROI-based method, amount of measures (features) in the connectivity profiles for the module-based method, and the amount of independent components in the ICA-based approach.

The voxel amount can be considered to be of highest - and significantly greater - magnitude, ranging from single- to triple-digit amounts of thousands of voxels per individual and/or experiment and depending on the resolution of the fcMRI data used for computations. The size of normative population involved, although for the five metrics in question varying extensively from 80 (as chosen by Nenning et al. [2]) to 1000 (as chosen by Stoecklein et al. [3]), may be for the purposes of our hypothesis estimated to range from hundreds to thousands, as higher normative sample sizes are vital for precise quantification of outliers in normative modeling [4], and thus positioned at the second magnitude rank. At magnitude rank 3 follows the amount of time-points within the individual fcMRI data files, which typically may range in the lower triple-digit amounts [5, 6]. Rank 4 may be allocated to the measure/feature amount of the module-based method (with precisely 91 measure reported as informing the index in [7]). On rank 5 follows the amount of ROIs involved in computation of the ROI-derived index (precisely 55 per hemisphere and functional connectivity topology [8] and, finally, the independent component amount for the ICA-based approach may be positioned at the magnitude rank 6, as Silvestri et al [9] reported analysis of 45 resting-state network components.

For rigor, this relation shall be formalized as

$$V \gg N \geq T \geq D \geq R \geq C \tag{2}$$

where $V$ - number of voxels; $N$ - number of individuals; $T$ - number of time-points; $D$ - number of connectivity features; $R$ - number of ROIs/modules/"nodes"; $C$ - number of relevant independent components.

It follows that an increment of growth of larger factors would contribute greater to the complexities influenced by these factors, and that products of larger factors or exponents thereof would result in a greater increase of complexities than products or exponents of smaller factors.

### 2 Computational Complexity Estimation of Algorithms

Here we put forth our estimates. For brevity, we omit the repetitive clarifications of formula element denominations, as those were given above.

#### 2.1 Naïve voxel-based algorithms

Both the Nenning (Nenning et al. [2]) and the dysconnectivity (Stoecklein et al. [3]) indices constitute naïve approaches - voxel-wise approaches that operate straightforwardly and with a stable performance irrespectively of the number of queries. With the prominent advantage of the significant accuracy at voxel-wise resolution, they possess, however, substantial resource requirements.

Making use of the Big O trivial property, one can estimate the algorithms for both indices to be in the order of complexity of the Pearson correlation coefficient computing operation, particularly at the moment of covariance matrix computation. Here, the most performance-heavy move consists in matrix multiplication. Computing both indices therefore has a time complexity of

$$\mathcal{O}(NTV^2) \tag{3}$$

From the aspect of space complexity, the same pipeline step also constitutes the moment of peak memory load. Therefore, the dynamic component of space complexity (i.e. the newly-generated products requiring storage) outweighs the static component (i.e. the initial data and necessary constants) and for both algorithms may be expressed as

$$\mathcal{O}(NV^2) \tag{4}$$

#### 2.2 Module-centric algorithm (n-PBSI)

The complexity estimation of the normative person-based similarity index [7] is based on the assumption that while Pearson correlation coefficients were used to assess the between- and within-module connectivity, the extent of computations, as reported by Doucet et al., was not uniform to the entire gray matter and concerned modules within and between networks, thus allowing us to assume "non-congestive", modular character of operations and disregard this step in search for the other resource-intensive operation. This allows us to assume the Spearman correlation coefficient computation during calculation of the n-PBSI for module cohesion and module integrations as the most resource-heavy part. With this in mind, the time complexity can be formulated as

$$\mathcal{O}(DN^2) \tag{5}$$

From the space complexity aspect, the dynamic component, consisting in newly generated feature vectors for every patient, remains quite light. Assuming streamlined feature computation, which implies dropping of Pearson-based connectivity maps immediately upon use as byproducts, it can be considered that the static component possesses greater weight. Therefore, the space complexity can be expressed as

$$\mathcal{O}(NVT) \tag{6}$$

#### 2.3 ICA-derivative algorithm (Spatial Similarity Index)

The network topography spatial similarity index [9] is markedly different from the other reported approaches due to dimensionality reduction, with the number of independent components employed in core analysis amounting to 45. Moreover, an informed estimation of both complexities is made challenging by employing MATLAB functions and algorithms, which permits only superficial assumptions. Given use of MATLAB, a degree of optimization may be considered inherent to Silvestri's method. Under these conditions, the time complexity may be assumed as

$$\mathcal{O}(NVTC) \tag{7}$$

Similarly to the previous approach, the assumption of strong baseline optimization allows to assume prevalence of static component over the dynamic component, letting us express the space complexity as

$$\mathcal{O}(NVT) \tag{8}$$

#### 2.4 Morgan Network Topology Method

The Morgan network topology method [8], as previously reported, employs ROI selection to form a functional connectivity topology and compute the results within it. Herein, the time complexity may be estimated as defined highest by the partial Pearson correlation computing for the purposes of analysis within the functional connectivity topology. In turn, this element is defined by edges - connections between ROIs, which the team around Morgan referred to as "nodes" in a later publication [10]. The amount of edges can be estimated as follows:

$$E = \frac{R(R-1)}{2} = \frac{R^2}{2} - \frac{R}{2} \tag{9}$$

With the assumption that $R = 55$ and $R^2 \gg R$, the time complexity of $\mathcal{O}(NET)$ can be represented through replacement of $E$ with $R^2$ as

$$\mathcal{O}(NR^2T) \tag{10}$$

From the space complexity aspect, the dynamic component once again remains light. The static component forms the bulk of the space complexity, represented as

$$\mathcal{O}(NVT) \tag{11}$$

### References


[1] Cormen, T. H., Leiserson, C. E., Rivest, R. L., & Stein, C. (2022b). Introduction to Algorithms, fourth edition. MIT Press.

[2] Nenning, K., Furtner, J., Kiesel, B., Schwartz, E., Roetzer, T., Fortelny, N., Bock, C., Grisold, A., Marko, M., Leutmezer, F., Liu, H., Golland, P.,

Stoecklein, S., Hainfellner, J. A., Kasprian, G., Prayer, D., Marosi, C., Widhalm, G., Woehrer, A., & Langs, G. (2020). Distributed changes of the functional connectome in patients with glioblastoma. Scientific Reports, 10(1). https://doi.org/10.1038/s41598-020-74726-1

[3] Stoecklein, V. M., Stoecklein, S., Galiè, F., Ren, J., Schmutzer, M., Unterrainer, M., Albert, N. L., Kreth, F., Thon, N., Liebig, T., Ertl-Wagner, B., Tonn, J., & Liu, H. (2020). Resting-state fMRI detects alterations in whole brain connectivity related to tumor biology in glioma patients. Neuro-Oncology, 22(9), 1388–1398. https://doi.org/10.1093/neuonc/noaa044

[4] Bozek, J., Griffanti, L., Lau, S., & Jenkinson, M. (2023). Normative models for neuroimaging markers: Impact of model selection, sample size and evaluation criteria. NeuroImage, 268, 119864. https://doi.org/10.1016/j.neuroimage.2023.119864

[5] Luckett, P. H., Park, K. Y., Lee, J. J., Lenze, E. J., Wetherell, J. L., Eyler, L. T., Snyder, A. Z., Ances, B. M., Shimony, J. S., & Leuthardt, E. C. (2023). Data-efficient resting-state functional magnetic resonance imaging brain mapping with deep learning. Journal of Neurosurgery, 139(5), 1258–1269. https://doi.org/10.3171/2023.3.jns2314

[6] Schmidt, T., Vannesjo, S. J., Sommer, S., & Nagy, Z. (2023). fMRI with whole-brain coverage, 75-ms temporal resolution and high SNR by combining HiHi reshuffling and multiband imaging. Magnetic Resonance Imaging, 103, 48–53. https://doi.org/10.1016/j.mri.2023.06.015

[7] Doucet, G. E., Glahn, D. C., & Frangou, S. (2020). Person-based similarity in brain structure and functional connectivity in bipolar disorder. Journal of Affective Disorders, 276, 38–44. https://doi.org/10.1016/j.jad.2020.06.041

[8] Morgan, V. L., Johnson, G. W., Cai, L. Y., Landman, B. A., Schilling, K. G., Englot, D. J., Rogers, B. P., & Chang, C. (2021). MRI network progression in mesial temporal lobe epilepsy related to healthy brain architecture. Network Neuroscience, 5(2), 434–450. https://doi.org/10.1162/netn_a_00184

[9] Silvestri, E., Moretto, M., Facchini, S., Castellaro, M., Anglani, M., Monai, E., D'Avella, D., Della Puppa, A., Cecchin, D., Bertoldo, A., & Corbetta, M. (2022). Widespread cortical functional disconnection in gliomas: an individual network mapping approach. Brain Communications, 4(2). https://doi.org/10.1093/braincomms/fcac082

[10] Morgan, V. L., Sainburg, L. E., Johnson, G. W., Janson, A., Levine, K. K., Rogers, B. P., Chang, C., & Englot, D. J. (2022). Presurgical temporal lobe epilepsy connectome fingerprint for seizure outcome prediction. Brain Communications, 4(3). https://doi.org/10.1093/braincomms/fcac128
